# Revisiting the impact of *Schistosoma mansoni* regulating mechanisms on transmission dynamics using SchiSTOP, a novel modelling framework

**DOI:** 10.1101/2024.02.16.24302940

**Authors:** Veronica Malizia, Sake J. de Vlas, Kit C.B. Roes, Federica Giardina

## Abstract

**Background:** The transmission cycle of *Schistosoma* is remarkably complex, including sexual reproduction in the human hosts and asexual reproduction in the intermediate host (freshwater snails). Patterns of rapid recrudescence after treatment and stable low transmission are often observed, hampering the achievement of control targets. Current mathematical models commonly assume regulation of transmission to occur at worm level through density-dependent egg production. However, conclusive evidence on this regulating mechanism is weak, especially for *S. mansoni*. In this study, we explore the interplay of different regulating mechanisms and their ability to explain observed patterns in *S. mansoni* epidemiology.

**Methodology/Principal findings:** We developed SchiSTOP: a hybrid stochastic agent-based and deterministic modelling framework to reproduce *S. mansoni* transmission in an age-structured human population. We implemented different models with regulating mechanisms at: i) worm-level (density-dependent egg production), ii) human-level (anti-reinfection immunity), and iii) snail-level (density-dependent snail dynamics). Additionally, we considered two functional choices for the age-specific exposure to water bodies. We compared the ability of each model to reproduce observed epidemiological patterns pre- and post-control, and we compared the successful models in their predictions of the impact of school-based and community-wide treatment.

Simulations confirmed that assuming at least one regulating mechanism is required to reproduce a stable endemic equilibrium. Snail-level regulation was necessary to explain stable low transmission. Only models combining snail- and human-level regulation with an age-exposure function informed with water contact data were successful in reproducing observed patterns. However, the predicted probability of reaching the control targets varied across successful models.

**Conclusions/Significance:** The choice of regulating mechanisms in schistosomiasis dynamics largely determines the model-predicted feasibility to reach control targets. Overall, the models that successfully explained observed patterns for *S. mansoni* suggest that reaching the control targets may be more challenging than currently thought. Conclusions highlight the importance of regulating mechanisms to be included in transmission models used for policy.

**Author Summary:** Schistosomiasis is a neglected tropical disease estimated to affect 200 million people worldwide and causes severe morbidity in endemic countries. The transmission of *Schistosoma* is complicated by the multiplication of parasites within freshwater snails that act as intermediate hosts. Rapid rebound of infection after mass treatment is commonly observed, as well as stable low transmission. Several mathematical models are currently employed to inform policy decisions, building on the predominant assumption of worm-level regulation through density-dependent egg production. We explored the interplay of different regulating mechanisms assumed at worm-, human-, and snail-level and their ability to explain observed epidemiological patterns. To this end, we developed SchiSTOP, a new modelling framework for *S. mansoni* transmission dynamics and considered 162 model variants. Results show that snail-level regulation is required to explain stable low transmission, while the combination of snail- and human-level regulation is necessary to reproduce a rapid rebound after control. We further show that the choice of regulating assumptions heavily influences conclusions about the feasibility to reach control targets. In particular, the models that successfully explained observed patterns for *S. mansoni* suggest that achieving control targets solely through mass drug administration may be more challenging than currently thought.

## Introduction

Schistosomiasis is a neglected tropical disease (NTD) caused by parasitic *Schistosoma* parasitic flatworm species. Widespread in tropical and sub-tropical regions, schistosomiasis is endemic in 78 countries with an estimated 236 million people requiring treatment in 2019 [1]. Responsible for severe morbidity such as complications due to chronic infections of the urinary tract or the intestine, schistosomiasis is considered the deadliest among NTDs [2]. The intestinal form of schistosomiasis is caused by *S. mansoni,* and urogenital schistosomiasis is caused by *S. haematobium* [3]. The transmission dynamics of *Schistosoma* between the human host and the water environment are peculiar and remarkably complex, due to two reproduction phases. The first-stage larvae (miracidia) multiply asexually in freshwater snails, acting as intermediate host, and develop in a second larval stage (cercariae) able to infect humans, where they mature, pair, and sexually reproduce. Monogamous in their nature, adult pairs of *Schistosoma* release eggs within the human host. Individuals with schistosomiasis contaminate freshwater sources with their excreta containing parasite eggs, which result in miracidia after hatching. The World Health Organization (WHO) 2030 Roadmap for NTDs delineates two control targets for schistosomiasis: elimination as a public health problem (EPHP) in all endemic countries and interruption of transmission (IOT) in 32% of the endemic countries [1]. EPHP is defined by WHO as achieving a prevalence of heavy intensity infections in school-aged children (SAC) below 1%. Infections are classified as heavy intensity if more than 400 eggs per gram of faeces (epg) are detected for *S. mansoni*, and >50 eggs per 10 mL urine for *S. haematobium*. IOT is defined by WHO as having no new autochthonous human cases in a defined geographical area.

Mass drug administration (MDA) to SAC with the anthelmintic drug praziquantel is the primary measure for the control of schistosomiasis. Praziquantel reduces the overall prevalence of infection in the population by killing adult worms within the human hosts [4]. However, the prevalence of infection typically rebounds to pre-control levels quickly after stopping MDA [1, 5, 6], but we currently lack a clear understanding of rebound pathways. Moreover, maintained transmission at low endemic levels (< 10% prevalence in SAC) has been observed [7–9], hampering the achievement of interruption of transmission. The WHO has recently provided a recommendation for additional control measures to prevent rebound of infection, including focal snail control and behavioural change interventions (through education and improvement of hygiene and sanitation standards) in endemic countries [10].

Mathematical models have historically been employed to reproduce and elucidate the transmission dynamics of *Schistosoma* species [11]. Deterministic models have been developed to describe the mean worm burden within humans over time [12–15], while more sophisticated agent-based models (ABMs) have included individual heterogeneities in transmission, exposure to infection and adherence to treatment [16–21].

Mathematical models for schistosomiasis require assumptions on mechanisms regulating transmission in order to reproduce a stable endemic equilibrium. In the absence of regulating mechanisms, the number of cercariae in the environment and the burden of worms in humans will grow unbounded, leading to an endemicity setting where the entire population is heavily infected. The assumption of density-dependence in egg production, according to which production of eggs by female worms diminishes with increasing adult worm burdens (density-dependent fecundity), has been largely accepted and employed with the rationale of overcrowding effects within the human host, in analogy with other intestinal parasites [13, 22]. However, scientific evidence for this assumption for schistosomiasis has been based on historic human autopsy data [23, 24], and the applicability of such data was disputed [25]. The availability of new molecular data analysed via sibship reconstruction [26] has provided some recent evidence of density-dependence in egg production for *S. haematobium* species, but the results were not conclusive for *S. mansoni,* where this question remains unresolved.

Regulating mechanisms at the level of the human host or at the level of the snail host, and the interplay of such mechanisms, have not been sufficiently included in current models used for policy. At the human level, host-acquired immunity has been suggested in antibodies detection studies [27–29] and was included in some deterministic age-structured models [30–32]. However, a clear and conclusive understanding of the processes of the development of acquired immunity (e.g. triggers, rate) and the extent to which it may restrain transmission is missing, especially for *S. mansoni* [33]. Modelling studies have shed some light on the role of the intermediate hosts in the transmission dynamics of schistosomiasis [34–36]. On the one hand, snails can itself regulate the transmission dynamics with an amplifying effect due to the multiplication of miracidia within the snail host. On the other hand, the snails constitute a potential restrain of transmission because of their competition for resources [36], the depletion of susceptible snails, and the increased snail mortality upon infection [37, 38]. Overall, insufficient attention has been given to describe both host-induced immunity and dynamics of the intermediate host in mathematical models employed for guiding policy decisions. There is a need to further understand the extent to which different regulating mechanisms occurring at worm-, human-, and snail-level can explain observed epidemiological patterns and how these affect the estimated impact of control interventions.

The aim of this study is to assess how well different modelling assumptions on the regulating mechanisms of schistosomiasis transmission dynamics can reproduce observed endemicity pre-control settings (low, moderate, high) and explain epidemiological patterns observed before, during and after MDA with praziquantel. We also investigate how such assumptions on regulating mechanisms and their interplay affect the feasibility to reach the control targets set by the WHO, in particular EPHP and IOT following two MDA strategies: targeting SAC or the whole community. In this work, we focused on *S. mansoni* species and developed SchiSTOP: a hybrid modelling framework for the transmission of *S. mansoni* between humans and snails that combines an agent-based model (ABM) describing the human and worm populations, and a deterministic compartmental model for the snail dynamics. We consider the combinations of three assumptions on the regulation occurring at worm level (density-dependence in egg production), human-level (anti-reinfection immunity), and snail-level (density-dependence in snails’ population growth).

## Methods

### Model structure

We developed SchiSTOP, an agent-based stochastic modelling framework for the transmission dynamics of schistosomiasis between the human hosts and the contaminated water environment via larvae multiplication in the intermediate host (freshwater snails). The dynamics of the snail population are included as an explicit deterministic compartmental model integrated into the ABM.

SchiSTOP consists of five main building blocks: the human population, the parasitic worms living in the human host, the two larval stages living in the contaminated water environment, and the snail population. The models include human, worm and snail population dynamics via births, aging, and deaths. For humans, migration is also considered. Infection in humans is then modelled through an age-specific exposure to water, potentially contaminated with cercariae. Maturation, reproduction, and death of worms within the human host are governed by stochastic events. Human hosts contribute to the contaminated water environment through excretion of eggs and thus release of miracidia, which can infect snails. Infection dynamics in snails bridge the steps between the asexual reproduction of miracidia in snails and the maturation into cercariae. Diagnostic schemes to assess infection in humans, and control interventions via 10 annual rounds of MDA are explicitly modelled, assuming 75% coverage [10]. We also take into account that a 5% of the population is systematically excluded from MDA, and we assume an imperfect drug efficacy of 86% killing rate of adult worms [39]. Detailed specifications of SchiSTOP and a full list of parameters are provided in the **S1 Text**.

SchiSTOP allows the implementation of different model variants (from here on called models) by varying assumptions on parameters and processes describing the dynamics of the disease. For the scope of this study, we considered and implemented 162 different models that reflect the 27 combinations of three regulating mechanisms acting at different levels of the transmission cycle (worm, human and snail) at three intensities (absent, mild, strong), as well as two different functions to model age-specific exposure to the water environment, and three endemicity levels (low, moderate, high). In particular, the three regulating mechanisms act at i) worm-level, through density-dependence in egg production, ii) human-level, through anti-reinfection immunity, iii) snail-level, through the density-dependent growth of snail population. The two functional choices for the age-specific exposure to the water environment are i) a model-based function derived from fitting an established schistosomiasis transmission model [21] to observed age-intensity profiles [39] (model-based), and ii) an alternative age-exposure function that we estimated from published water contact data [40, 41] (water-contacts-based). To inform the latter, we extracted frequency of water contacts adjusted for body surface and time of day of contact reported for three age groups (0-9, 10-19, 20+ years old) from an extensive data collection on direct water contact observations in a village in Northern Senegal [40]. To refine the modelling of the exposure in the last age-group (20+), various age-exposure profiles from Kenya [41] were used. Both functions are implemented as a relative exposure and displayed in **Fig 1A**.

**Fig 1.**
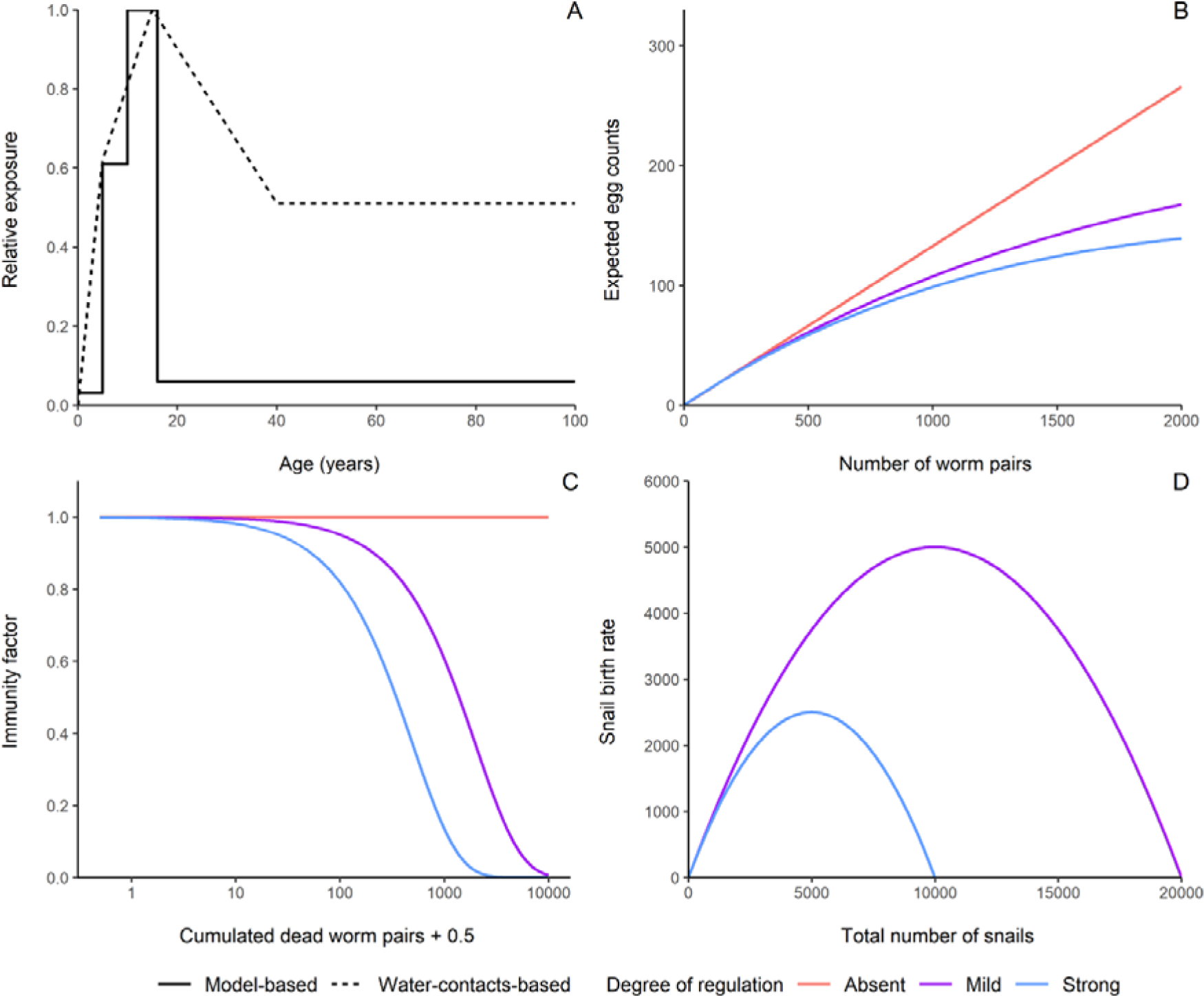
Modelling assumptions. **(A) Age-exposure functions employed in the models.** The functions describe the relative exposure by age of the human host, using 1.0 for the highest value (i.e., at age 15). Both functions are interpolations of reported data points: the solid line is a piece-wise constant function representing the relative exposures derived from literature (model-based exposure function) at 0, 5, 10, and 16+ years of age. The dashed line is a piece-wise linear function based on the relative exposures extracted from published water contacts data at 0, 5, 15, and 40+ years of age. **(B) Worm-level regulation.** The expected egg counts are function of the number of mature worms, which are modelled as pairs within the human host. The mean number of eggs per sample a is set at 0.13 (Absent), 0.14 (Mild), or 0.18 (Strong). The density-dependence parameter *z* is set at 0 (Absent), 0.0007 (Mild), or 0.004 (Strong). **(C) Human-level regulation.** The immunity factor is displayed as a function of cumulated dead worms, presented on a logarithmic scale. The immunity coefficient *α_imm_* is 0 (Absent), 0.0005 (Mild), or 0.002 (Strong). **(D) Snail-level regulation.** The snail birth rate, following a logistic growth, is displayed as a function of the total number of snails. The snail carrying capacity K is set at 20000 (Mild), or 10000 (Strong). When no regulation at the snail-level is assumed, the model does not include explicit snail dynamics, therefore the corresponding degree is not displayed.

### Worm-level regulation: density-dependent egg production

We consider mature worms to live in pairs within the human host. For each human individual,, the relationship between the expected excreted egg load (*ε_i_*) and the number of carried adult worm pairs (*wp_i_*), is implemented as an exponential saturating function, describing density-dependent fecundity of female worms as follows: *ε*_i_ = *αwp_i_ e^-zfwi^*. In this function, *fw_i_* = *wp_i_* /2 approximates the female worm load, the parameter *α* represents the maximum fecundity of female worms (i.e., the expected egg produced per worm pair in absence of density-dependence) and *z* is a parameter dictating the level of saturation in egg production. The worm-level regulating mechanism for different values of the parameters *α* and *z* is displayed in **Fig 1B**. The values of *α* and *z* were set to numbers available from previous work or tuned to produce a mild or strong regulation. To do so, we first set *z* to 0 and assign *α* the value from literature [42] in absence of worm-level regulation, assuming the excreted egg load (*eggs_i_*) to linearly depend on the number of adult worm pairs. Secondly, for parametrising a strong worm-level regulation, we set *z* from literature [17] and tuned *α* to reproduce the same initial growth as in absence of density-dependence. Finally, we tuned both parameters for a mild worm-level regulation.

### Human-level regulation: anti-reinfection immunity

The human immunological response, which develops naturally, is assumed to be induced by the death of adult worms, as proposed in published immunological studies [33, 43]. Immunity protects the host from reinfection by hindering the maturation of cercariae into adult worms, and the amount of this immunity increases as a function of the cumulative number of died adult worms. The model describes this behaviour via an immunity factor given by exp(–*α_imm_ dwp*), which reduces the rate of parasite acquisition by the human host. Here, *α_imm_* is the immunity parameter and *dwp* is the individual cumulated dead worm burden, defined in terms of worm pairs and inclusive of worms undergone both natural mortality and mortality upon MDA. We do not consider decay of immunity within the human lifespan. The human-level regulating mechanism for different values of *α_imm_* is displayed in **Fig 1C**. The immunity factor is multiplied times the exposure rate, which determines the force of infection acting on humans (*FOI_h_*). In absence of anti-reinfection immunity, we set *α_imm_* to 0, implying that the past exposure to infection (and therefore death of worms) does not affect the chance to acquire new parasites. As a reference for a strong human-level regulation, we extracted *α_imm_* from literature [32]. We set the parameter to a lower value to define a mild degree of regulation.

### Snail-level regulation: density-dependent population growth

The snail phase of the parasite life cycle is explicitly modelled through an equation-based framework and embedded into each ABM. We employed a deterministic compartmental model to simulate births, deaths, exposure, and infection of snails. Susceptible snails *S(t)* reproduce according to a logistic growth with birth rate *β(t)* due to competition for resources:

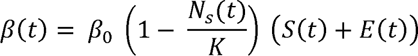

Here, *β_0_* is the maximum reproduction rate in absence of competition [36], *N_s_(t)* the total snail population size, and *K* the carrying capacity. The birth rate of snails, dictating the snail-level regulating mechanism, is displayed in **Fig 1D**. for different values of *K*. The carrying capacity *K* was tuned to parametrise a mild or strong degree of the snail-level regulating mechanism.

Susceptible snails can be invaded by miracidia present in the water environment after egg excretion by human hosts and become exposed (*E(t)*). The force of infection acting on snails *FOI_s_* is proportional to the abundance of miracidia, through a coefficient *η* representing the transmission parameter on snails. Miracidia live in exposed snails for a period during which they mature and asexually reproduce into cercariae. Susceptible and exposed snails undergo natural mortality, according to a mortality rate *v*[36]. When the parasites are mature, the snails become infected *I(t)* at a rate *τ* equal to the inverse of their assumed maturation period [17, 44]. Infection on snails causes an increased mortality (_νI_) and castration [36]. Therefore, only susceptible and exposed snails contribute to reproduction (see formulation of *β(t)*). Infected snails shed cercariae *C(t)* into the environment, according to a cercarial per capita production rate *λ* [36]. The abundance of shed cercariae determines the force of infection acting on humans *FOI_h_*. Cercariae naturally die with a rate *γ* before infecting humans. The dynamics of this module are thus described by the following system of ordinary differential equations.

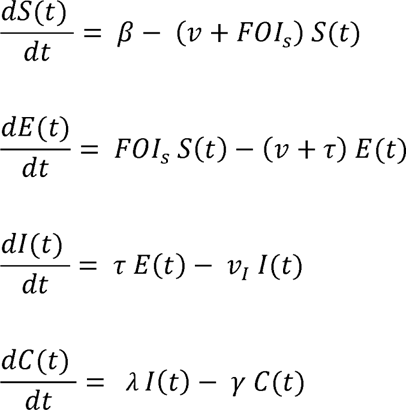

When there is no regulation at the snail-level, we assume that the cercarial uptake by the environment at time *t* coincides with the number of miracidia excreted at the previous time step, which coincides with the maturation period into the intermediate host (1 month). The full list of parameters and the specifications of the snails’ infection dynamics can be found in **S1 Text**, Intermediate host.

### Simulations

Three parameters, namely the overall transmission parameter on humans *ζ*, the level of worm aggregation *k_w_,* and the transmission parameter on snails η, were varied within a grid search framework with the aim to initiate all models at the same setting of either low, moderate, or high endemicity. The initial settings were defined on the basis of three prevalence indicators and their targets informed from literature: the infection prevalence in SAC, as per WHO definition (average of 10%, 30%, 60%, in low, moderate, and high endemicity, respectively) [10], the prevalence of heavy infections in SAC (average of 0%, 6%, 20%) [45], and prevalence of infection in snails (average of 6% in all settings) [37]. First, a visual inspection of the three prevalence indicators allowed refining the search. The prevalence of infection in snails is mostly influenced by the parameter η, about which very little information is available. We therefore swept the parameter η until the models reproduced a prevalence in snails close enough to the target value (6%). Afterwards, the grid search for the three transmission parameters was performed in the reduced space and the parameters’ values minimising the root mean squared error (RMSE) between the predicted and desired targets, were chosen for each model. The RMSE was computed combining and scaling the three target indicators with most of the weight assigned to the prevalence of any infection in SAC, which was allowed to vary about ± 0.1% with respect to the desired threshold. The file with selected parameters for each model and the full code is publicly available at https://github.com/VeronicaMalizia/SchiSTOP. SchiSTOP was implemented and run using R programming language 4.2.2 [46].

After the selection of parameters, we ran 100 stochastic simulations for each model to capture inherent models’ variability and used the average trends for comparison. We assessed and compared the ability of each model to explain three observable patterns: i) the age-intensity profiles expressed as mean detected eggs per gram (epg) of faeces as a function of age, ii) the effect of 10 annual rounds of MDA administered to SAC, iii) the rebound of prevalence after treatment. The number of eggs per gram of faeces (epg) is expressed as geometric mean across age groups (see **S1 Text,** Human Demography) and stochastic realizations of the models. Although geometric means lead to biased estimators of the true mean, they have traditionally been used in the analysis of data from field studies [47]. We defined models as successful when they could i) reproduce a convex age-intensity curve with a typical peak of intensity of infection in adolescents, ii) bring the prevalence down of at least 20% of their pre-control levels after 10 years of MDA, and iii) result in a rebound of prevalence to values close to pre-control levels, after stopping MDA, even in low endemicity settings [1, 48-50]. After stopping MDA, we expect that the prevalence of infection in SAC should go back to pre-control levels within 20 years. In order to parametrise the model and define the above-mentioned criteria for model plausibility, we undertook an expert elicitation process that involved multiple steps. We first identified experts in the field ranging from parasitology, epidemiology, experimental and field research. All experts had a strong publication record, practical experience, and a deep understanding of the disease dynamics. All experts were informed about the purpose of the expert elicitation process, the goals of our research, and the specific areas in which we sought expertise. In particular, we conducted structured interviews and one workshop, all organized within the SchiSTOP project [51]. We asked questions related to 1) the role of the intermediate host and how to interpret malacological surveys, 2) exposure and mechanisms of immunity in terms of triggers, protection, and decay and 3) prevalence patterns before and after control.

Lastly, we used the successful models to compare the predicted probabilities (based on 100 repeated simulation runs) to meet the control targets of EPHP and IOT, considering two different treatment strategies: 10 rounds of annual MDA administered to SAC (school-based), and 10 rounds of annual MDA administered to all individuals older than 2 (community-wide), as per the most recent WHO guidelines towards the 2030 targets [10]. EPHP is reached when the prevalence of heavy intensity of infections in SAC is below 1%, and IOT when the prevalence of any intensity of infection in SAC is 0.0%. We define a stochastic run to have met the target if the prevalence indicator is still below the control threshold 50 years since the end of the MDA programme.

## Results

### Reproducing initial settings

The defined initial settings of low, moderate, and high endemicity were reproducible only with a subset of models (147 out of 162). Clearly, the presence of at least one regulating mechanism is required to reproduce a stable endemic equilibrium for all three endemicity settings. In addition, adopting the model-based age-exposure function, worm- and human-level regulation alone was not sufficient to reproduce a stable low endemic equilibrium (≤ 10% prevalence of any infection in SAC). Explicit snail population dynamics at any intensity of regulation is essential for reaching low stable endemic equilibria. With the alternative definition of age-exposure function based on water contacts we found that any regulating mechanism can initiate the model in a low endemicity setting. In particular, snail-level regulation allows very low endemic equilibria.

When the system does not reach the required endemic equilibrium, the number of simulated cercariae in the environment and the average worm load in humans can either grow without upper limits, leading quickly to an endemicity setting where the entire population is infected, or die out, leading to a disease-free situation. Models without any regulating mechanism and models without snail-level regulation combined with the model-based exposure function are not successful in reproducing stable equilibria for all endemic settings and have therefore been excluded from the assessment of further results. Only exception to this exclusion is the model based on strong worm-level regulation alone and model-based exposure function. Although this model does not allow to generate steady initial settings with low endemicity, it is included for reference in further comparisons, as current models for policy use the same assumptions.

### Reproducing observed epidemiological patterns

#### Age-intensity profiles

The assumptions on human-level regulation via immunity and on the age-exposure functions have the largest impact on the resulting shape of the age-intensity profiles, whereas varying assumptions on regulation occurring at worm- and snail-level has a minor effect. **Fig 2** compares the age-intensity profiles for different degrees of regulation in humans (immunity, by colours) and both age-exposure functions, for all three endemicity settings. The figure shows that the peak in intensity of infection reproduced by all models is followed by i) a steep drop for adults with the model-based exposure function, and ii) a smoother decline in adults leading to higher egg counts, using the exposure function based on water contacts. In the latter, implementing human-level regulation from absent to strong, shifts the peak from 20 to 10 years old on average and considerably decreases the mean intensity in adults. Interestingly, despite a relatively high exposure in adults assumed by the water contacts-based exposure function, a strong regulation in humans can explain the low intensity of infection in adults (20+) that is sometimes observed in specific exposure settings (**Fig 2D** and **2F**). The effect of immunity on the intensity of infection is not seen in low endemicities, because the accumulation of dead parasites in human hosts is too low to trigger an immune response.

**Fig 2.**
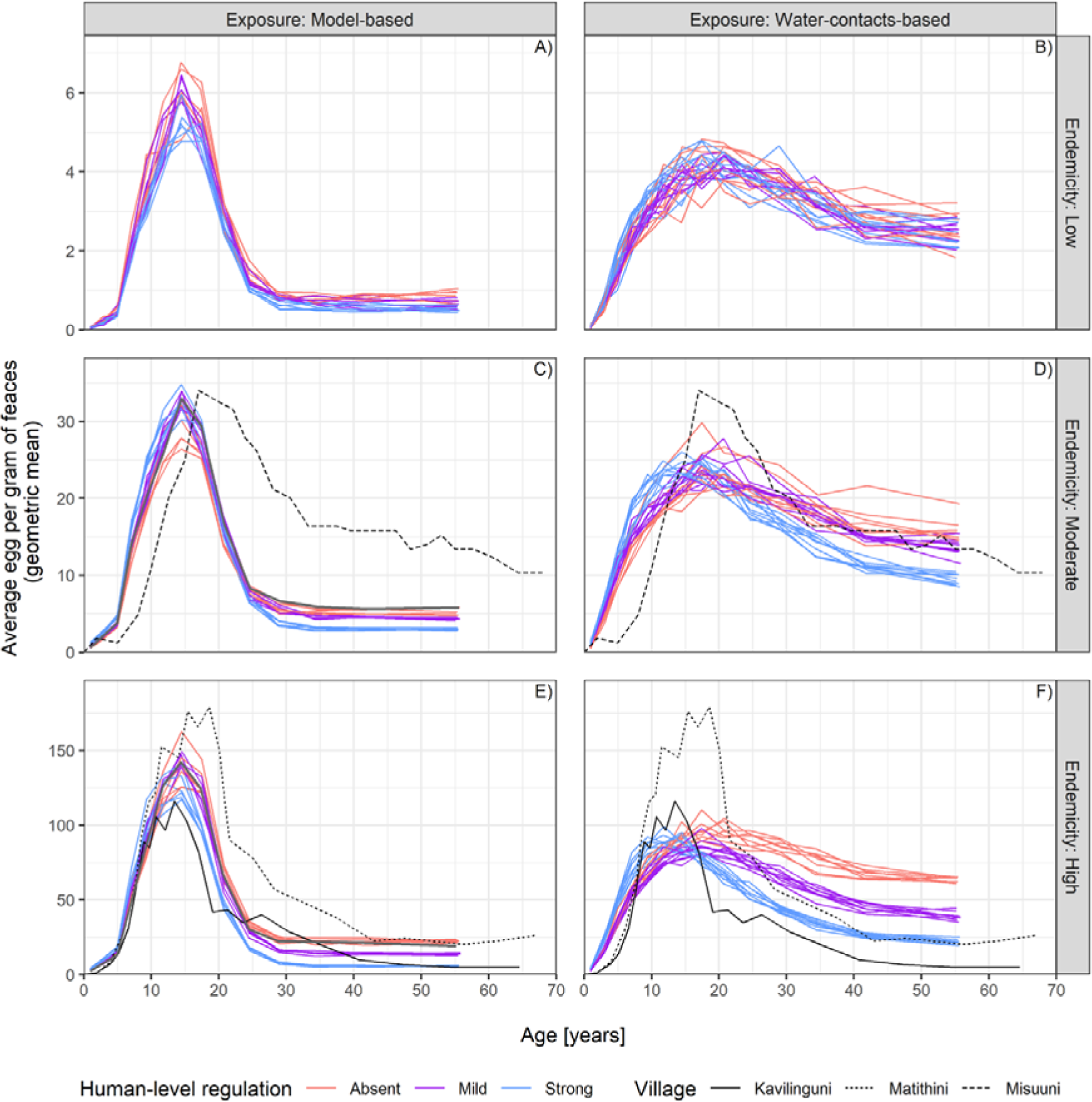
Age-intensity profiles by varying assumptions on age-exposure and immunity mechanism. For each choice of the age-exposure function (columns) and different endemicity settings (rows), a single panel shows the simulated egg counts on the y-axis (mean epg is displayed, over age group and 100 stochastic realizations of the models) by age on the x-axis. The age-intensity profiles are displayed at pre-control, for all models that could successfully reproduce all three pre-control endemicity settings. Each line depicts results from a single model. The degree of regulation assumed at human level via immunity is highlighted with different colours. Lines of the same colour are due to combinations (3×3) of worm- and snail-level regulation, given the same choice of immunity. Age bins are defined as to have the same number of individuals in each bin. The grey line in the left-hand-panels (C and E) represents a model with strong worm-level regulation only and the model-based exposure function. This model does not allow to generate stable initial settings with low endemicity, but it is included here for reference, as current models for policy use the same assumptions. The black lines indicate age-intensity profiles observed in three Kenyan villages and published elsewhere [52].

Observed age-intensity profiles from three Kenyan villages published elsewhere [52] are also plotted in **Fig 2** (black lines) for comparison with simulated profiles. It can be seen that the model-based exposure function allows to reproduce high endemicity settings, but the water contacts-based exposure function requires a degree of regulation in humans to reproduce low intensity in adults. This is different in moderate endemicity settings, where the effect of the model-based exposure function on the age-intensity profiles is likely to be too strong at older ages, with respect to the alternative exposure function.

The age-intensity profiles disaggregated for all models are displayed in **S1 Appendix.**

#### Effectiveness of MDA and prevalence bounce-back

All models reproduce similar decreasing trends of prevalence during MDA (**Fig 3**). Assumptions on human-level regulation and the chosen age-exposure function are the main determinants of the drop in SAC prevalence due to treatment, depending on the endemicity setting. With a model-based exposure function, MDA has a stronger effect on prevalence in SAC. The prevalence drops to lower levels after 5 MDA rounds if compared to the results obtained by using the exposure function based on water contacts. This can be explained by the fact that the simulated control strategy targets the SAC group, where most of the exposure occurs, particularly in the model-based exposure function. A strong regulation in humans also predicts a more effective treatment with respect to assuming absent or mild regulation. In fact, MDA itself contributes to the development of anti-reinfection immunity by its acting mechanisms that kills adult worms [29].

**Fig 3.**
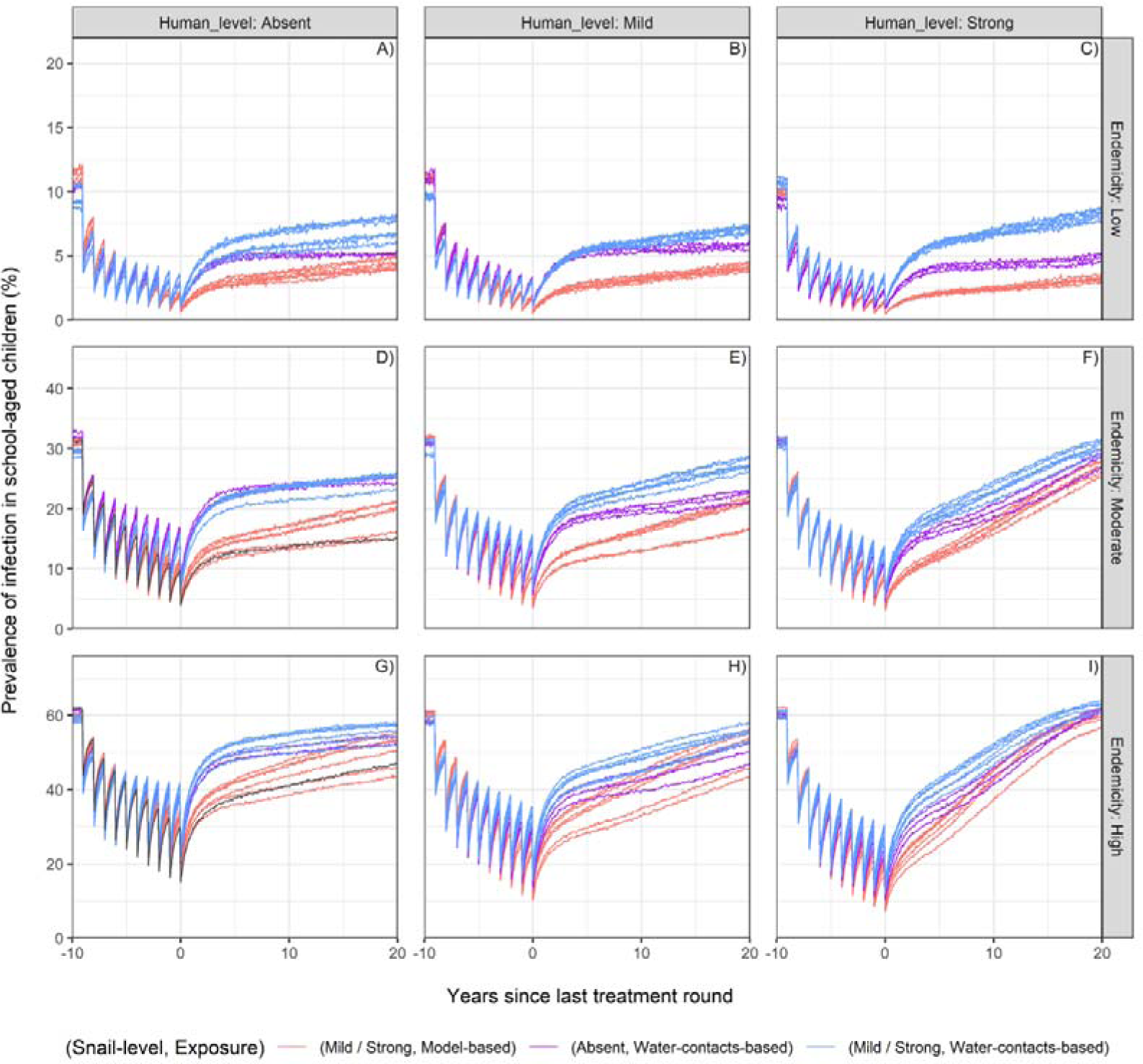
Prevalence timelines in SAC during and after treatment. For different degree of human-level regulation via immunity (columns) and different endemicity settings (rows), single panels show the infection prevalence in school-aged children on the y-axis (mean of 100 stochastic realizations of the model) by years since the last treatment round on the x-axis, for all models that could successfully reproduce all three pre-control endemicity settings. Each line depicts results from a single model. Lines of the same colour represent groups of models as indicated in the legend. The grey line in panels D) and G) represents a model with a strong worm-level regulation only and the model-derived exposure function. The model is excluded from the analyses because it does not allow stable initial settings with low endemicity, but it is added here for reference as current models for policy use the same assumptions. For all models, treatment is annually administered to 5-15 years old individuals with a coverage of 75%, 5% of target population systematically untreated, and a drug efficacy of 86%.

The ability of each model to reproduce the natural prevalence bounce-back is further shown in **Fig 3**, by the prevalence trends up until 20 years after the last MDA round. The figure shows that the combination of the age-exposure function based on water contacts and a mild or strong degree of regulation in snails is essential to reproduce a bounce-back of prevalence close to pre-control levels in all endemicity settings, within 20 years after termination of the MDA programme. The rationale for this result lies in a reduced impact of treatment as a consequence of the water contact-based exposure function, as previously explained, in addition to higher values of transmission parameters required if considering snail-level regulation. By assuming a strong regulation at human level, the bounce-back is much more prominent, although the prevalence drops to lower values during treatment with respect to assuming the regulation to be absent or mild. In fact, assuming strong regulation at human level requires higher transmission parameters in order to reproduce the initial endemicity settings. The models found successful in explaining a plausible rebound of prevalence after treatment are not significantly influenced by the degree of worm-level regulation considered.

The timelines of SAC prevalence during and after the MDA programme disaggregated for all models are displayed in **S2 Appendix.** Overall, the models that were successful in explaining observed patterns (blue lines in second and third columns in **Fig 3**), both in terms of pre-control age-intensity profiles and bounce-back of prevalence after treatment were those that combined an age-exposure function based on water contacts with a mild or strong degree of regulation at both snail- and human-level.

### Feasibility to reach the control targets

Despite reproducing similar epidemiological patterns, successful models can still differ in their predictions of probability to reach the control targets. Results that follow include only such selected models, with the exception of the model based on only strong worm-level regulation and model-based exposure function, as current models for policy use the same assumptions.

#### Reaching control targets with 10-year annual MDA to school-aged children

We found that all models agree that treating SAC only with 10 years of annual MDA will not allow to reach the EPHP in high or moderate endemicity settings. In low endemicities, we start from a setting where the prevalence of high intensity infection in SAC is already below 10% (EPHP has already been met). Here, the target of interest is the IOT and none of the models predict a positive probability to reach this target despite 10 rounds of annual MDA to SAC.

#### The impact of community-wide annual MDA

Expanding the 10-year annual MDA programme to all individuals older than 2 years of age, leads to EPHP in low-endemicity settings for all successful models. The probability to reach EPHP in moderate endemicity settings turns to a range varying from 58% to 100% in all models with a mild human-level regulation (**Fig 4**). The mechanism of anti-reinfection immunity (human-level regulation) has the highest impact on the probability to reach EPHP: a strong human-level regulation decreases the probability to a range between 15% and 28% combined with another regulating mechanism (snail or worm) (**Fig 4**). In fact, a strong human-level regulation is associated with higher transmission and larger heterogeneity of worms (input files with transmission parameters can be found in the public repository https://github.com/VeronicaMalizia/SchiSTOP) in the human hosts to reproduce the initial endemic setting. In high endemicity settings, it is still feasible to reach EPHP with a probability ranging between 2% – 46% if human-level regulation is mild. The probability shows a decreasing trend if mild to strong worm-level regulation is assumed. In fact, worm-level regulation mechanism decreases the impact of MDA as individuals with high worm loads will still have a similar egg output after treatment, therefore keeping their contribution to transmission. Consequently, prevalence rebounds earlier than in models without worm-level regulation, when a linear relationship between worm burdens and expected egg counts is assumed. According to all other models, the probability to reach EPHP is close to zero in high endemicity settings (**Fig 4**).

**Fig 4.**
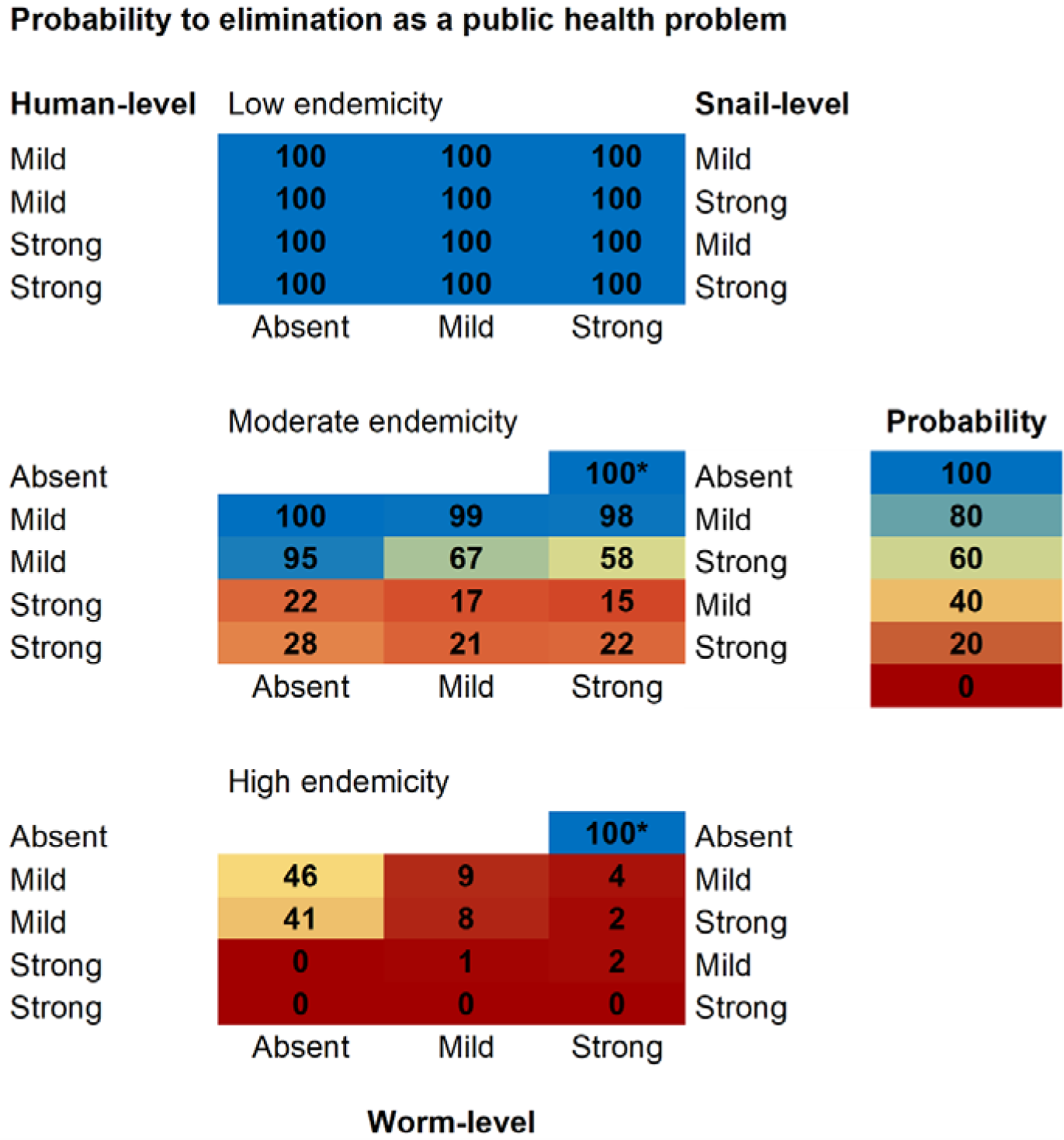
Probability to reach elimination as a public health problem with a 10-year annual MDA to all individuals older than 2. The figure reports predictions for the probability (portion of stochastic runs monitored at 50 years after end of treatment) of reaching elimination as a public health problem (EPHP, prevalence of heavy infections in school-aged children < 10%), across models successful in reproducing all observed patterns for schistosomiasis. Successful models assume an age-exposure function based on water contacts. Regulating mechanisms can be at human-level (left side, via immunity), snail-level (right side, via density-dependence in population growth), and worm-level (bottom, via density-dependence in egg production). *This model assumes a strong worm-level regulation only and the model-derived age-exposure function. It was not successful in reproducing observed patterns within our modelling framework but is added here for reference as current models for policy use the same assumptions. For all models, treatment is annually administered to 2+ years old individuals with a 75% coverage, 5% of target population systematically untreated, and a drug efficacy of 86%.

Community-wide MDA also increases the chances of reaching IOT with respect of only treating SAC, even in moderate (15 – 41 %) and high (0 – 7 %) endemicity settings (**Fig 5**). The probability of interrupting transmission in low endemicity settings jumps from 0% (if treating only SAC) to a range of 88 – 98% across the different models. The probability to reach both control targets was found to be higher when using assumptions as employed in current models used for policy (marked by *).

**Fig 5.**
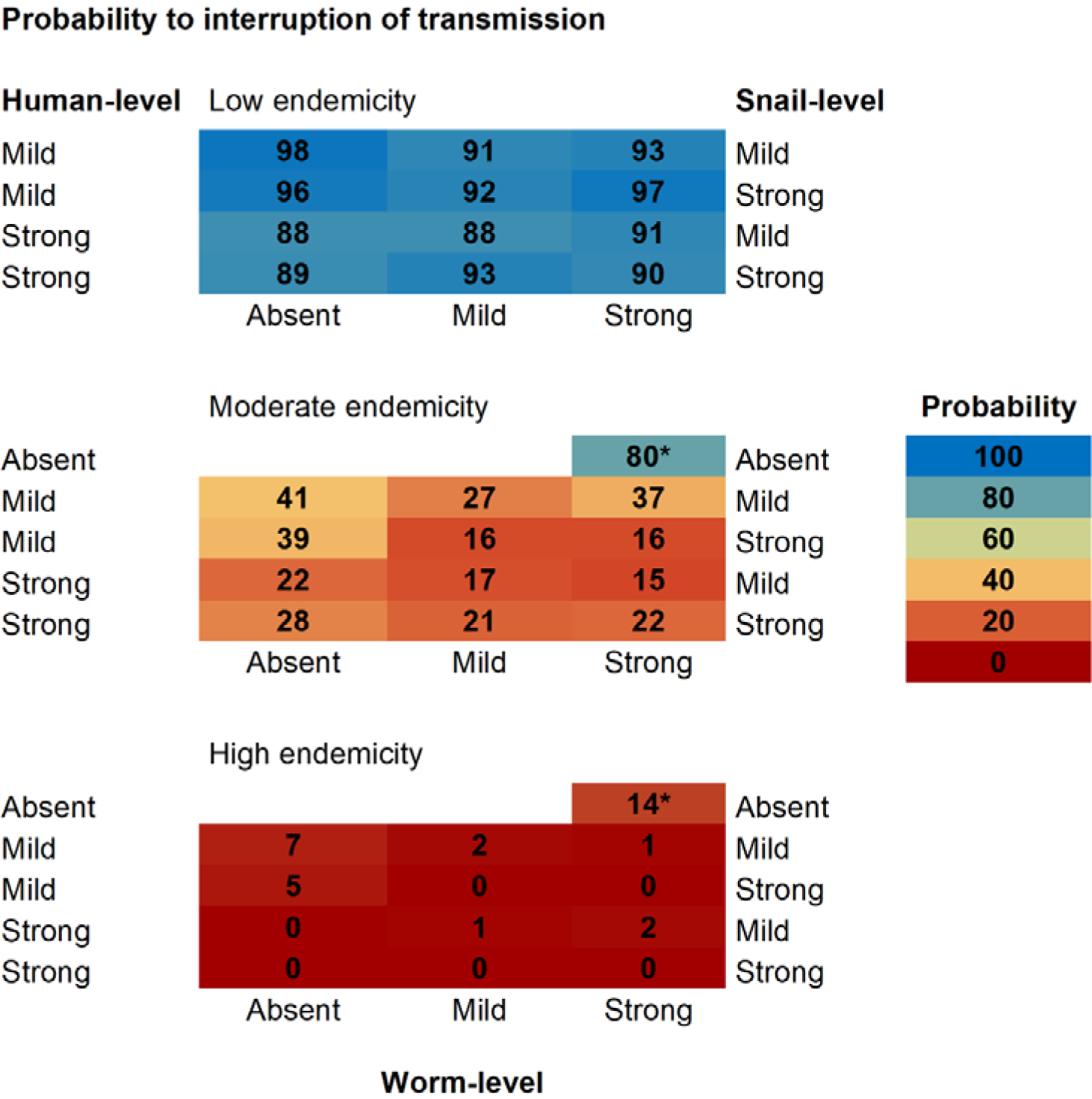
Probability to reach interruption of transmission by 10-year annual MDA to all individuals older than 2. The figure reports predictions for the probability (portion of stochastic runs monitored at 50 years after end of treatment) of reaching interruption of transmission (0% prevalence of any infection in school-aged children), across models successful in reproducing all observed patterns for schistosomiasis. Successful models assume an age-exposure function based on water contacts. Regulating mechanisms can be at human-level (left side, via immunity), snail-level (right side, via density-dependence in population growth), and worm-level (bottom, via density-dependence in egg production). * This model assumes a strong worm-level regulation only and the model-derived age-exposure function. It was not successful in reproducing observed patterns within our modelling framework, but it is added here for reference as current models for policy use the same assumptions. For all models, treatment is annually administered to 2+ years old individuals with a 75% coverage, 5% of target population systematically untreated, and a drug efficacy of 86%.

## Discussion

The objective of our study was to revisit the modelling assumptions on the regulating mechanisms for the transmission of *S. mansoni* and to investigate the impact of such assumptions on the feasibility to reach the WHO control targets. To do so, we developed SchiSTOP, a new modelling framework for the transmission dynamics of *S. mansoni*. SchiSTOP was used to run a large number of simulations, varying the assumptions on the regulating mechanisms and the age-exposure to infection.

We found that not all combinations of regulating mechanisms and age-exposure function can successfully reproduce the defined pre-control endemicity settings (low, moderate, high). In particular, without regulating mechanisms, the system leads quickly to settings where the entire population is highly infected, as previously demonstrated in other modelling studies [53]. Moreover, stable low endemic equilibria (prevalence of infection in SAC ≥ 10%) cannot be reproduced with the modelling assumptions on age-exposure and only worm-level regulation that are considered in current models used for policy. To our knowledge, there are no schistosomiasis modelling studies in the literature that aim to focus on low transmission settings. We found that snail-level regulation is crucial to reproduce low stable endemic equilibria, within SchiSTOP. In fact, snail dynamics with a varying carrying capacity can stabilise the system providing a continuous reservoir of infection even at very low transmission levels in humans, thanks to the force of infection acting on snails which contributes to maintain transmission. Overall, our results show that a degree of regulation in snails and in humans combined with the water contacts-based exposure function is needed to reproduce expected age-intensity profiles and a plausible rebound of prevalence after termination of an MDA programme.

The assumptions made on the regulating mechanisms are particularly relevant when the models are used for policy recommendations. In fact, the predicted probabilities to reach the control targets differ across successful models, and they are overall lower compared to the predictions obtained using the regulation assumptions of current models used for policy recommendations. Results show that according to all successful models, it is unlikely to meet EPHP and IOT when treating SAC only. If MDA is expanded to community-wide treatment of all individuals older than 2, the feasibility increases, even regarding IOT in moderate and high endemicity. This is in line with previous studies [54].

The present work also highlights the importance of informing the age-exposure function with water contact data. In fact, the model-based age-exposure function only performs well in reproducing age-intensity profiles in high endemicity settings, while the drop in intensity of infection at older ages may be too strong in lower endemicities. This is because this function was derived from fitting a transmission model solely considering regulation at worm level to the age-intensity profiles from different African villages, all characterized by high endemicity [11, 15, 17, 55, 56]. Available data show that age-intensity profiles, although generally peaking in 10-20 years old, can appear differently in older ages describing heterogeneities in endemicity, exposure, and regional-specific settings [47]. The alternative age-exposure function based on water contacts has the ability to reproduce the variety in age-intensity profiles, with immunity leading to a peak shift to lower ages and lower intensity in adults [57]. Here we show that some degree of human-level regulation via immunity is likely to occur in order to explain observed age-intensity relationships in high endemicity settings, using the water contacts-based exposure function. A recently published review provided evidence for water contacts being robustly associated with infection in both children and adults, across heterogeneous settings. These findings [58] shed light on the definition of at-risk populations and therefore in need of treatment and highlight the need of standardised tools for exposure measurements. However, how to properly define and quantify exposure to infection on the basis of observable data such as number of contacts with contaminated water, duration of contact and area of the body exposed, remains a challenge.

Overall, the modelling assumptions commonly employed in schistosomiasis modelling and used for informing control strategies (i.e., in terms of age-exposure and only worm-level regulation) are found not adequate to reproduce low endemic pre-control settings and rebound of prevalence after MDA, within SchiSTOP. Predictions to reach the control targets under those assumptions are generally more optimistic compared to the same predictions for the successful models in our study. This is because i) with a model-based function, the MDA has a stronger effect on prevalence in SAC, ii) the assumption of a strong worm-level regulation requires lower transmission parameters and a higher heterogeneity of worms to represent moderate and high endemicity scenarios, compared to those required by other regulating mechanisms.

Our modelling framework, SchiSTOP, has some limitations, mainly related to the limited knowledge and reliability of data available to more precisely parametrise regulating mechanisms. In particular, we assumed that acquired immunity leads to a protective effect towards repeated infections, as suggested for *S. mansoni* [33]. However, the long-term dynamics of immunity still constitute a research and data gap [33]. While the current understanding points towards a slow development of immunity triggered by the death of adult worms (both naturally and by praziquantel therapy) [59], not much is known about the duration of such protection. We chose not to account for a decay of anti-reinfection immunity for the scope of these analyses. In addition, some studies suggest that the main effect of acquired immunity is a decrease in worm fecundity. This has been proposed for *S. haematobium* [60, 61], but does not seem to be the case for *S. mansoni* [33, 62]. The assessment of infection in snails is also subject to uncertainty due to sampling procedures, and diagnostic methods to assess infection. Field and laboratory data have shown that the detected prevalence of infected snails (thus shedding cercariae) is usually quite low [37, 63-66], but exhaustive explanation of such low prevalence is lacking and no evidence of correlation between infection rates in snails and humans has been identified. We chose a fixed 6% prevalence of infection in snails to define all three endemicity initial settings. Molecular tools and genetic data are further necessary to deepen our understanding of variations in susceptibility to schistosome species by specific intermediate host snails.

Furthermore, we compute probability of IOT as number of stochastic runs that meet the target up to 50 years after termination of the MDA programme, despite monitoring and evaluation in the field are likely to occur within shorter time intervals [10], to ensure capturing pathways of late recrudescence. A rebound after elimination, as well as maintained low transmission, can nevertheless occur via other mechanisms like movement of infected individuals [67] or cercariae/snail movement through water stream. SchiSTOP does not take this into account, and we consider it to play a minor role for schistosomiasis, given its focal nature [3].

In this work, SchiSTOP has been used to revisit the assumptions of regulating mechanisms for *S. mansoni* transmission. However, its formulation is suitable to answer diverse research questions about the epidemiology and control of schistosomiasis. For instance, the presence of a specific module for the dynamics in snails allows for a thorough assessment of the impact of snail control interventions.

In conclusion, the present work highlights the importance of considering regulating mechanisms at different levels of the transmission cycle in models for schistosomiasis transmission and control, as they largely determine the predicted impact of interventions. We showed that some degree of regulation in both snail- and human-level is required to explain observed epidemiological patterns typical for schistosomiasis. Our findings support the current WHO guidelines, recommending community-wide treatment to all individuals > 2 years old. This control strategy greatly enhances the probability to reach the control targets, also in high endemicity settings. However, the probability to reach these targets is likely not as high as predicted by the currently used models that only assume worm-level regulation. We stress on the need for a better understanding of the mechanisms regulating transmission and persistence of schistosomiasis in endemic settings.

## Supporting information

**S1 Text. Full specification of SchiSTOP and complete list of parameters employed for the analyses.**

**S1 Appendix. Age-intensity profiles for the complete set of models.** For each choice of the age-exposure function and each endemicity setting (titles), single panels refer to a fixed combination of the assumptions of worm-level regulation (columns) and snail-level regulation (rows). A single panel shows the simulated egg counts on the y-axis (mean epg is displayed, over age group and 100 stochastic realizations of the model) by age on the x-axis. The age-intensity profiles are displayed at stable pre-control settings, by varying the degree of regulating mechanism in humans (colours). Age bins are defined as to all be equally sized.

**S2 Appendix. Effectiveness of treatment and prevalence bounce-back for the complete set of models.** For each choice of the age-exposure function and endemicity setting (titles), single panels refer to a fixed combination of the assumptions of human-level regulation (columns) and snail-level regulation (rows). A single panel shows the infection prevalence in school-aged children on the y-axis (mean of 100 stochastic realizations of the model, single runs as shaded lines) by round of treatment on the x-axis. The prevalence timelines are displayed by varying the degree of worm-level regulation (colours). For all models, treatment is annually administered to 5-15 years old individuals with 10 repeated rounds, a coverage of 75%, 5% of target population systematically untreated, and a drug efficacy of 86%.

## Data Availability

The study does not make use of collected and reported field data.

## Acknowledgments

The authors gratefully acknowledge the experts who contributed to the inspiring panel discussions organized within the SchiSTOP project. The discussions guided the modelling decisions about regulating mechanisms and interpretation of observed patterns for *S. mansoni*. The participants: Professor Poppy Lamberton (School of Biodiversity, One Health and Veterinary Medicine, Wellcome Centre for Integrative Parasitology, University of Glasgow, Glasgow, UK), Dr Stefanie Knopp (Swiss Tropical and Public Health Institute, Basel, Switzerland), Professor David J Civitello (Department of Biology, Emory University, Atlanta, GA, USA), Professor Russell J Stothard (Centre for Tropical and Infectious Diseases, Liverpool School of Tropical Medicine, Liverpool, UK), Dr Antonio Montresor (Department of Control of Neglected Tropical Diseases, World Health Organization, Geneva, Switzerland), Dr Maurice Odiere (Neglected Tropical Diseases Unit, Kenya Medical Research Institute, Centre for Global Health Research, Kisumu), Professor Meta Roestenberg (Department of Parasitology, Leiden University Medical Center, Leiden, The Netherlands), Dr Govert J van Dam (Department of Parasitology, Leiden University Medical Center, Leiden, The Netherlands), Dr Jean T Coulibaly (University of Felix Houphouet-Boigny, Abidjan, Côte d’Ivoire).

