## Supplementary material for "Revisiting the impact of *Schistosoma mansoni* regulating mechanisms on transmission dynamics using SchiSTOP, a novel modelling framework": S1 Appendix

### Age-intensity profiles

**Age-intensity profiles for the complete set of modelling scenarios.** For each choice of the age-exposure function and each endemicity setting (titles), single panels refer to a fixed combination of the assumptions of worm-level regulation (columns) and snail-level regulation (rows). A single panel shows the simulated egg counts on the y-axis (mean epg is displayed, over age group and 100 stochastic realizations of the model) by age on the x-axis. The age-intensity profiles are displayed at pre-control, by varying the degree of regulating mechanism in humans (colours). Age bins are defined as to all be equally sized.

#### 1. Assumption for the age-exposure function: "Model-based

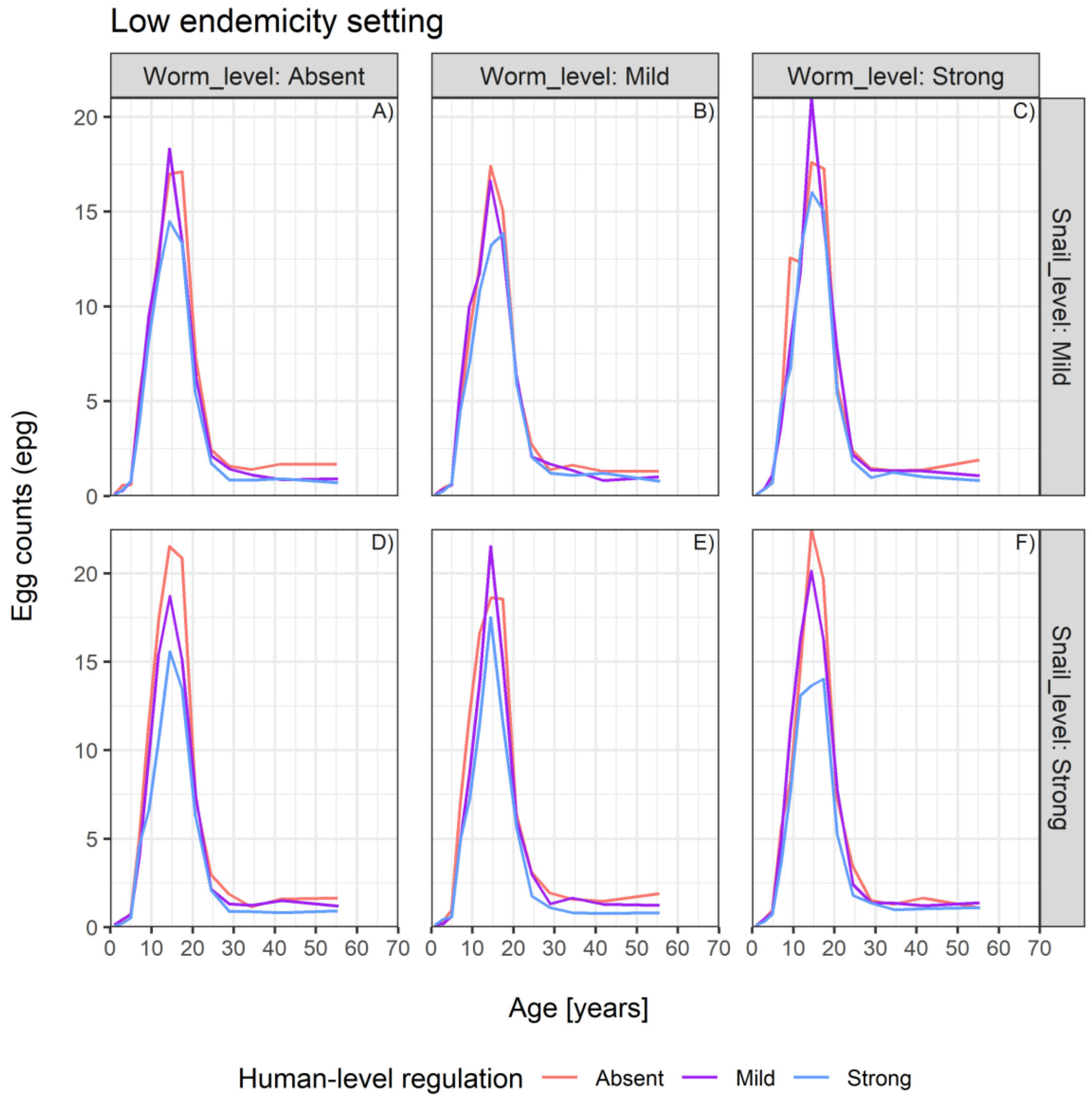

Low endemicity setting
Worm_level: Absent
Worm_level: Mild
Worm_level: Strong
Snail_level: Mild
Snail_level: Strong
A)
B)
C)
D)
E)
F)
Egg counts (epg)
Age [years]
Human-level regulation
Absent
Mild
Strong

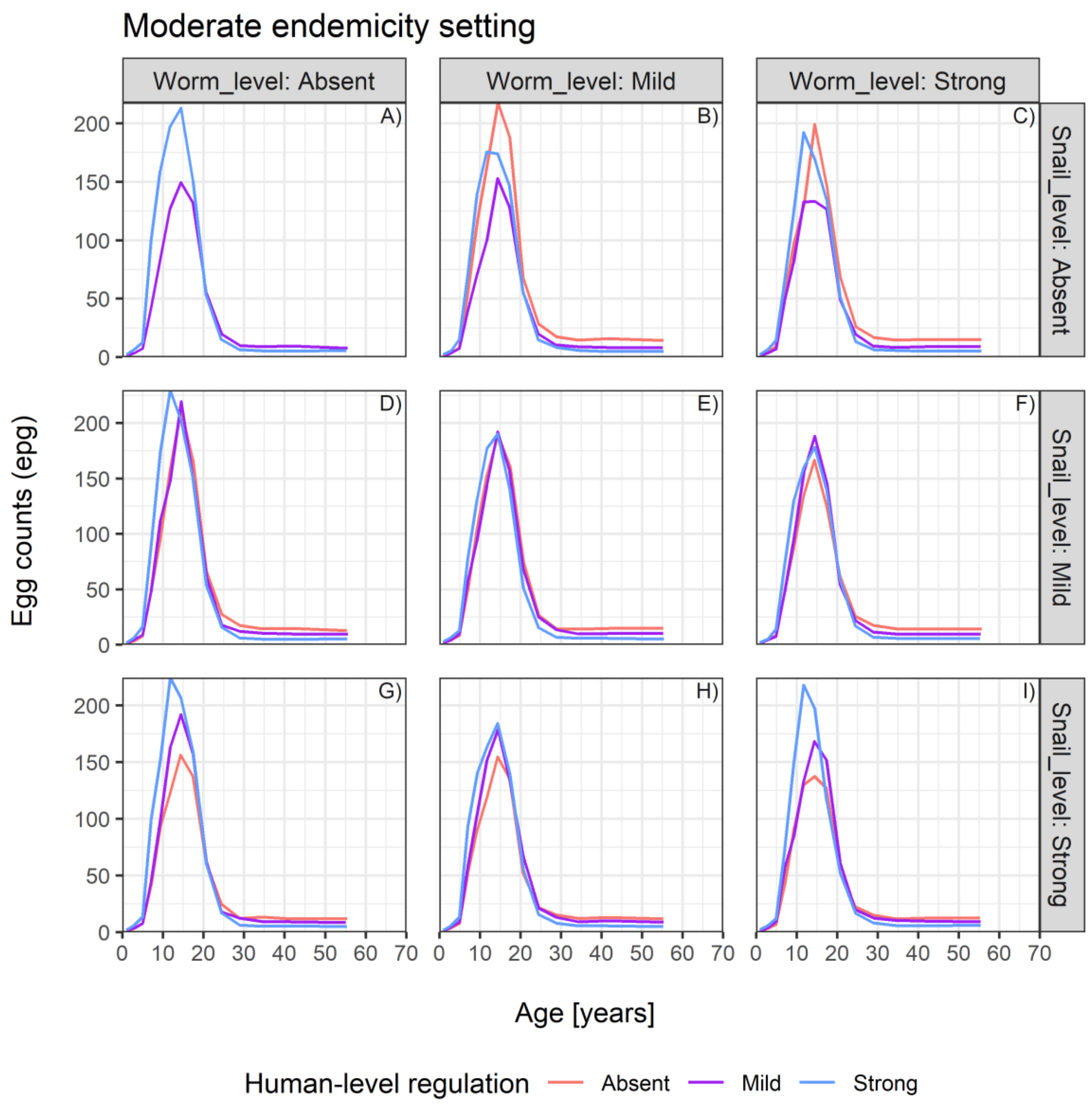
Moderate endemicity setting
Worm_level: Absent
Worm_level: Mild
Worm_level: Strong
Snail_level: Absent
Snail_level: Mild
Snail_level: Strong
A)
B)
C)
D)
E)
F)
G)
H)
I)
Egg counts (epg)
0
50
100
150
200
Age [years]
0
10
20
30
40
50
60
70
Human-level regulation
Absent
Mild
Strong

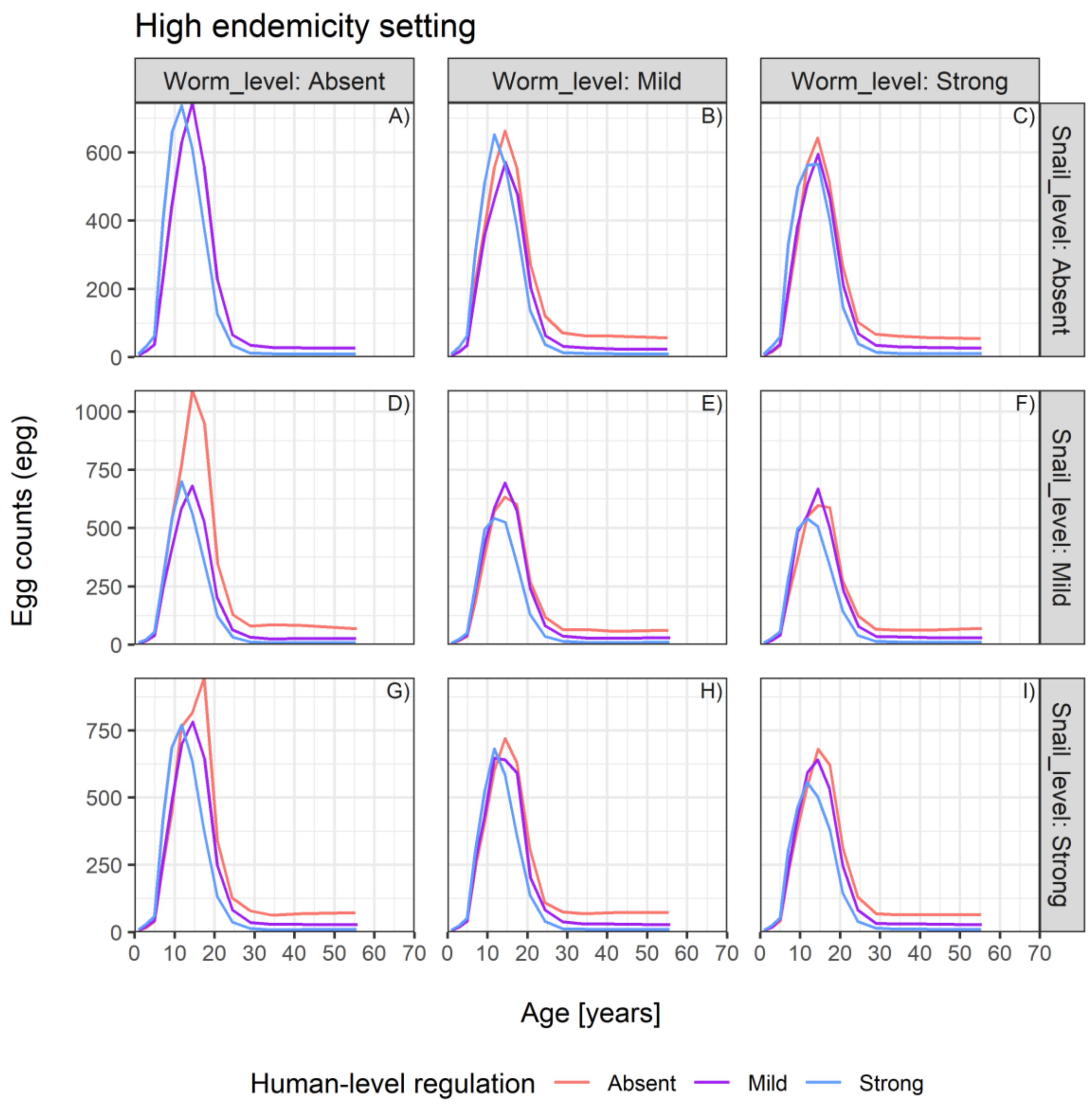
High endemicity setting
Worm_level: Absent
Worm_level: Mild
Worm_level: Strong
Snail_level: Absent
Snail_level: Mild
Snail_level: Strong
A)
B)
C)
D)
E)
F)
G)
H)
I)
Egg counts (epg)
0
200
400
600
0
250
500
750
1000
0
250
500
750
0 10 20 30 40 50 60 70
Age [years]
Human-level regulation
Absent
Mild
Strong

**2. Assumption for the age-exposure function: “Based on water contacts”**

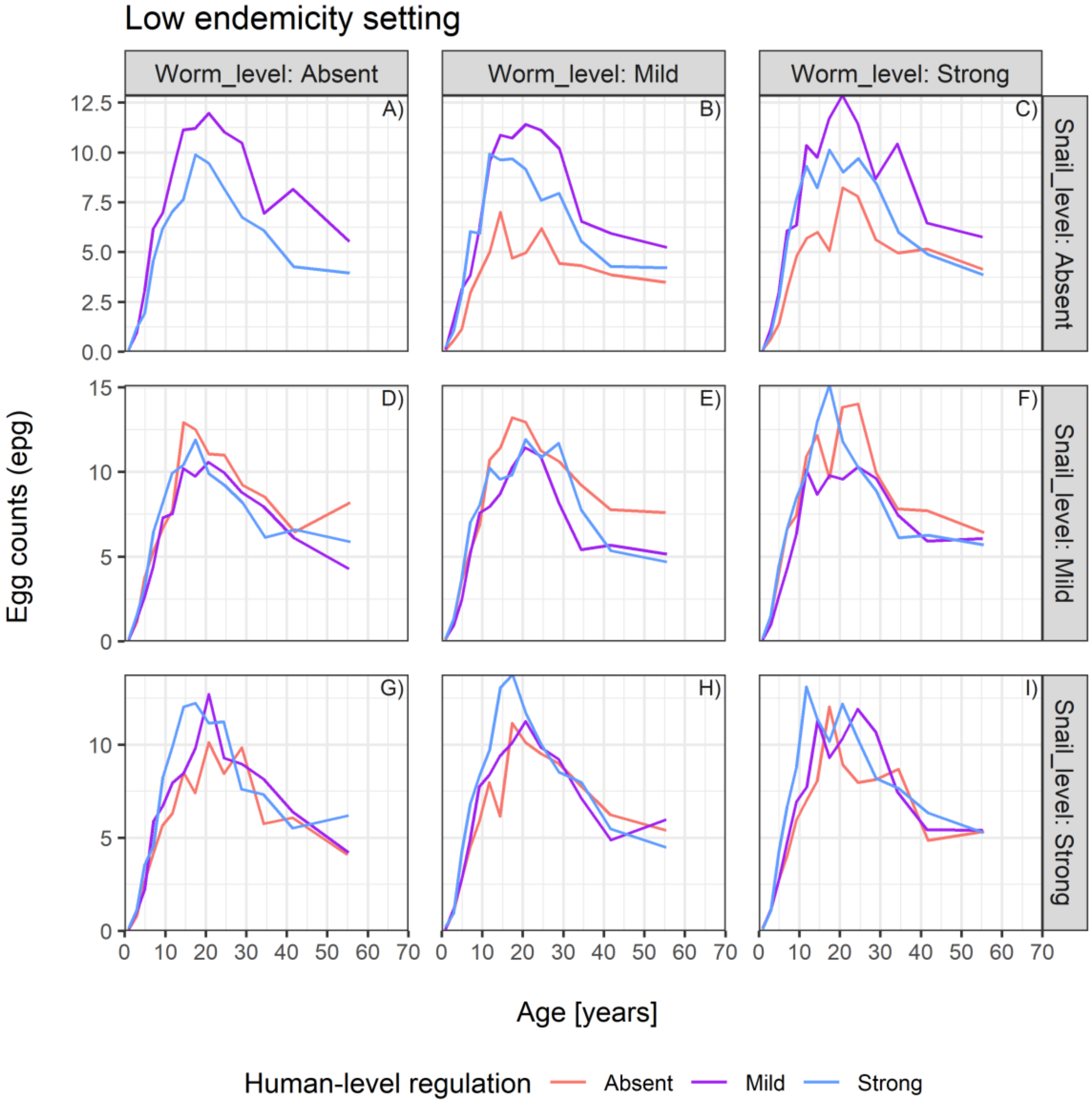

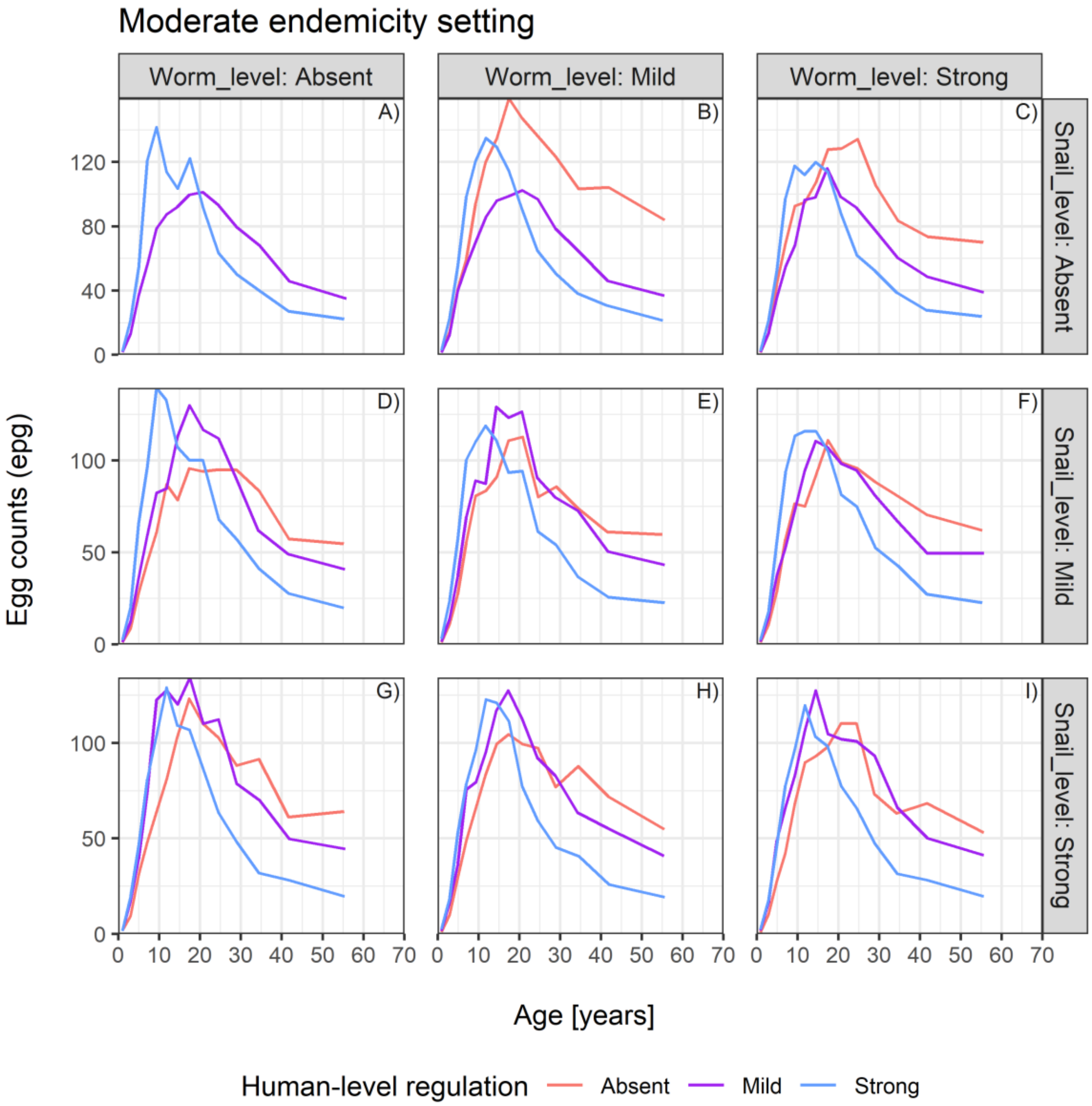

Moderate endemicity setting
Worm_level: Absent
Worm_level: Mild
Worm_level: Strong
Snail_level: Absent
Snail_level: Mild
Snail_level: Strong
A)
B)
C)
D)
E)
F)
G)
H)
I)
Egg counts (epg)
Age [years]
0 10 20 30 40 50 60 70
0
40
80
120
0
50
100
Human-level regulation
Absent
Mild
Strong

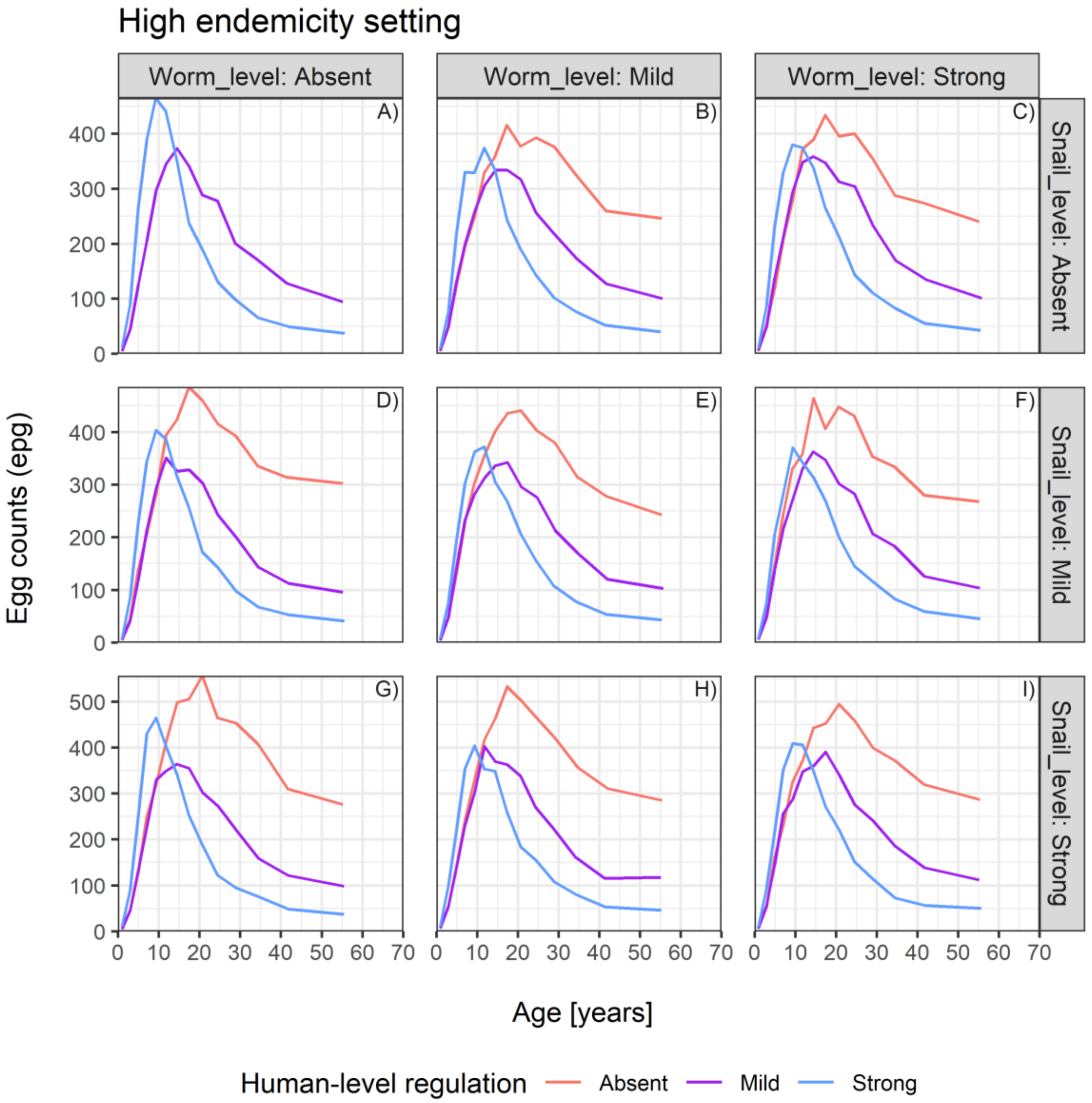
High endemicity setting
Worm_level: Absent
Worm_level: Mild
Worm_level: Strong
Snail_level: Absent
Snail_level: Mild
Snail_level: Strong
A)
B)
C)
D)
E)
F)
G)
H)
I)
Egg counts (epg)
Age [years]
Human-level regulation
Absent
Mild
Strong
