## Supplementary material for "Revisiting the impact of *Schistosoma mansoni* regulating mechanisms on transmission dynamics using SchiSTOP, a novel modelling framework": S1 Text

#### Introduction

This document contains full specification of the modelling framework SchiSTOP, including functions and parameters employed for the simulations. We implemented an agent-based stochastic model (ABM) to reproduce the transmission dynamics of schistosomiasis between the human hosts and the contaminated water environment, via larvae multiplication in the intermediate host, namely freshwater snails. The dynamics of the snail population are explicitly included via a deterministic model consisting of a system of ordinary differential equations (ODEs), integrated into the ABM. The five main building blocks of SchiSTOP are: the human population, the parasitic worms living in the human host, the two parasitic larval stages living in the contaminated water environment, and the snail population.

The ABM is based on stochastic events updated at discrete time steps $t$ of 1 month. The ODEs system is defined at continuous time scale $\tilde{(t)}$ and events are governed by daily rates, considering the short lived larval stages of *Schistosoma mansoni.* At each time step $t$ of the ABM, the ODEs system is solved in a time horizon $h$ equal to 1 month. Afterwards, the solutions of the ODEs system serve as initial conditions for the next ABM-time step $t + 1$.

The model is written in R programming language, version 4.2.2 [1] and the code is available for consultation and downloading at the public online repository `https://github.com/VeronicaMalizia/SchiSTOP`.

Here on, we refer the subscript $i$ to the $i$-th human individual alive at time $t$. The human population at each time step $t$ is tracked as a matrix $N \times m$ where $N$ is the number of alive individuals (i.e., the current population size) and m the columns storing the following individual characteristics: age, sex, individual susceptibility to infection, parasite acquisition rate, number of juvenile worms, number of adult worm pairs, cumulative dead worm pairs, and observed egg counts. All parameters employed in the model and mentioned in the next sections of this document are listed and documented in Table 1.

### Human demography

The human population is governed by births, deaths, aging and migration. The human demography is parameterised to follow the age-distribution in Uganda as reported by the Ugandan bureau of statistics [2].

The number of newborn individuals is deterministically obtained with a fixed birth rate $rate_b$. People at birth have zero worm/egg loads and their sex is assigned as random draw with probability 0.5.

Individuals migrate according to a fixed monthly net migration rate $rate_m$, which is tuned to reach an equilibrium population size of $N = 1000$, assuming the model reproduces the transmission dynamics in a rural community. Individuals of age between 5 and 55 years are eligible for migration.

Each month individuals can die according to an age- and sex- specific death probability. In case of death, the individual is removed from the population matrix.

The population is accordingly updated and aged of a period equal to $t$. The demographic dynamics are run to the equilibrium for 200 years and the final age distribution is used to initialise the transmission model (Fig 1).

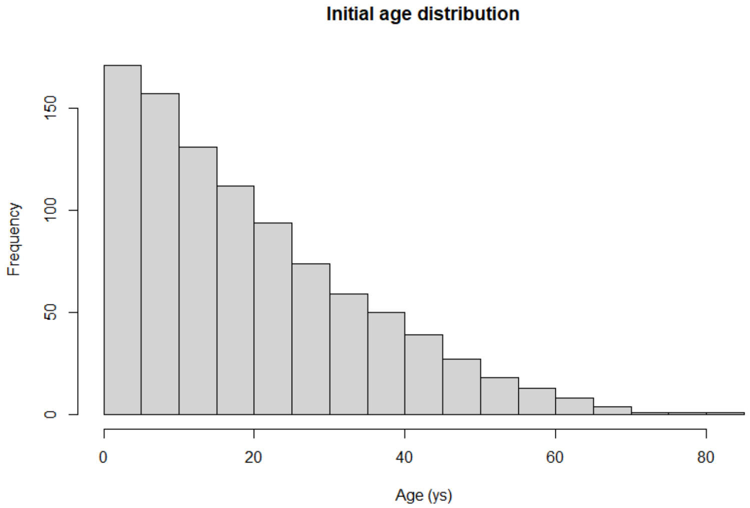


**Fig 1. Initial age distribution.** The histogram represents the age distribution used to initialise the population in the transmission model. This distribution is extracted after running the demographic dynamics to the equilibrium, for a total of 200 years. The human demography is parameterised to follow the age-distribution of Uganda as estimated by the Ugandan bureau of statistics.

### Infection dynamics

The transmission cycle of *S. mansoni* occurs between the human host and the contaminated water environment, mediated by freshwater snails acting as intermediate host. The first-stage larvae (miracidia) multiply in freshwater snails and develop in a second larval stage (cercariae) able to infect humans upon exposure. Individuals infected with *S. mansoni* contribute to the water environment with their excreta containing parasite eggs, thereby maintaining transmission.

#### Human exposure to infection

The force of infection acting on each human host is defined as follows:

$$FOI_{H_i}(t) = \phi_i(t)\ \exp(-\alpha_{imm} dwp_i(t))$$

where $\phi_i(t)$ describes the parasite acquisition rate, multiplied by an exponential factor describing the effect of human-level regulation implemented as anti-reinfection immunity: this dictates the level of protection from consecutive reinfections. The immunity coefficient $\alpha_{imm}$ is function of the dead adult worm pairs accumulated up to the current time step $(dwp_i(t))$. The value assumed for the immunity coefficient $\alpha_{imm}$ modulates the degree of the regulating mechanism in humans.

The parasite acquisition rate $\phi$ in each human host is defined as a function of the current amount of cercariae in the environment $(C(t))$, the transmission parameter $\zeta$ and the relative exposure to infection $Ex_i$:

$$\phi_i(t) = \frac{C(t)\ \zeta\ Ex_i}{\sum_{i=1}^{N} Ex_i}$$

The individual relative exposure to infection is $Ex_i = Exp\,(a_i)\,is_i$, where:

$Exp\,(a_i)$ is the relative exposure of an individual with age $a$, defined as a piece-wise function of user-defined exposure rates for a finite set of ages. This incorporates the age-specific pattern of exposure to the contaminated water;

$is_i$: individual susceptibility to infection, which captures personal factors influencing the chance to get exposed, e.g., contacts with water due to occupation. This parameter is assumed to follow a gamma distribution with mean 1.0 and shape and rate (or 1/scale) equal to $k_w$. The individual susceptibility of a person is assigned at birth and remains constant throughout lifetime.

Subsequently, each human host acquires new juvenile worms $(jw_i(t))$ according to a Poisson process with a rate given by the $FOI_{H_i}(t)$. The process of worms acquisition is therefore a Poisson-Gamma mixture, namely the individual worm burden follows a negative binomial distribution with aggregation parameter $k_w$. Such distribution has been shown to be adequate for describing the overdispersion of worms around the mean [3].

Newly acquired worms are assumed to be juveniles for a given pre-patent period. Juvenile worms do not mate nor reproduce. Mortality of juvenile worms is neglected. Once the pre-patent period is over, juvenile worms are randomly assigned sex according to a fixed probability of 0.5 and considered mature and patent to pair and reproduce.

#### Worm life within the human host

In the model, mature worms live in pairs. Individual worm pairs $(wp_i(t))$ are defined as the minimum number between female and male juvenile worms (all possible pairs are formed). Worms which do not pair will not survive the next time step. In the rest of the transmission cycle, mature worm pairs are considered as infective units for simplicity, supported by the monogamous nature of *Schistosoma*. Similarly to the assumptions used by previous models [4], the adult worms' lifespans follow an Erlang distribution with a rate $\lambda$ and a shape equal to a specified number of stages throughout which the worm pairs are distributed within each human host. The newly paired worms enter the first stage and are subsequently aged through all the remaining stages before they die. In each stage, the portion of worm pairs aging to the next step is given by $\psi = e^{-\lambda} = e^{-(s/Tw)}$ assuming exponential survival within each stage. Here, $s$ is the number of considered stages and $Tw$ the adult worm lifespan.

Worm pairs are expected to reproduce and to pass eggs into the human intestine. The eggs will be released into the environment through the faeces. The model accounts for egg detection via the Kato-Katz diagnostic test, that uses stool smear samples of 41.7 mg for egg counting. Assuming that all paired worms reproduce within a time step of one month, we define the individual expected number of eggs observed in a stool sample as: $\mu_i\,(t) = \ \alpha\ wp_i(t)\ e^{-z\ \frac{wp_i(t)}{2}}$ . Here, $wp_i(t)/2$ approximates the current female worm load, the parameter $\alpha$ represents the fecundity parameter (i.e. the expected number of eggs / worm pair / stool sample, in absence of density dependence in egg production) and $z$ is a density-dependency factor regulating the level of saturation in egg production, and therefore tuning the regulating mechanism at worm level. In a modelling setting without regulation occurring at worm level (via density dependence in egg production), we set $z = 0$ assuming the produced egg load to linearly depend on the number of worm pairs.

#### Diagnostic test

SchiSTOP simulates results of a given number of repeated Kato-Katz diagnostic tests, at pre-defined moments along the simulation. The result of the diagnostic test is drawn from a negative binomial distribution with mean equal to $\mu_i(t)$ at the current time step, and aggregation parameter $k_e$. If multiple tests are taken from each simulated individual, the results are averaged per person.

#### Human contribution to the water environment

The number of eggs ($eggs_i$) by which each individual contributes to the water environment is given by multiplying $\mu_i$ by the daily average of excreted quantity of faeces. We define the individual contributions ($co_i$) as:

$$co_i\,(t) = eggs_i\,(t)\;\; Con(a_i)$$

where $Con(a_i)$ indicates the relative contribution of an individual with age $a$, defined as a piece-wise function of user-defined contribution rates, for a finite set of ages. This accounts for any age-specific pattern in human contribution to infection.

We assume that all excreted eggs hatch in the water and mature into miracidia in a negligible simulation time. We can therefore immediately update the total miracidial uptake into the water environment $M(t)$ as the sum of individual contributions:

$$M\,(t) = \sum_{i=1}^{N} co_i(t)$$

### Intermediate host

#### Implicit intermediate snail host

In the simplest model formulation, no regulating mechanism is assumed to act at snail level. In such model variant, the dynamics in the intermediate host are implicitly modelled as maturation of miracidia into cercariae. The miracidia released in the water environment will simply mature into cercariae in a period $h$ which approximates the maturation period within the intermediate host, i.e., $C\,(t) = M\,(t - h)$.

#### Explicit intermediate snail host

SchiSTOP allows explicit modelling of the intermediate host, by means of a separate module for the simulation of infection dynamics within the snail population. The module consists of a deterministic equation-based model that represents the role of snails and their interaction with the human population.

The deterministic module is implemented as a compartmental model (SEIC) [5]

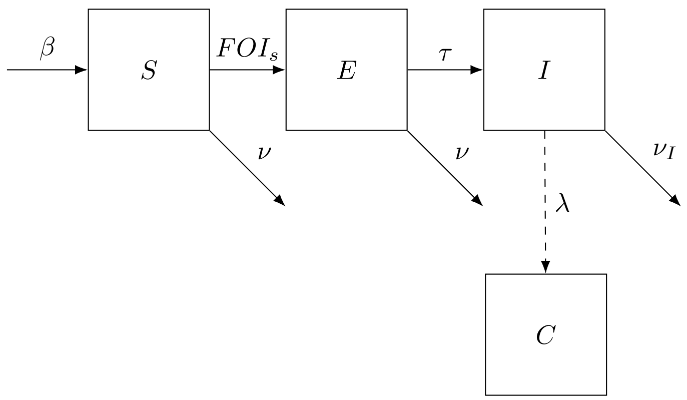


**Fig 2. Compartmental module simulating the transmission dynamics in the snail population and the cercarial production.**

where births, deaths, exposure and infection of snails occur. Finally, cercariae are shed in the water environment. The course of events is governed by daily rates. See Fig2 for a schematic representation of the snail transmission dynamics. The state of the system $(S(t'),\ E(t'),\ I(t'),\ C(t'))$ describes in order: the amount of susceptible, exposed, infected snails, and cercariae available at ODE-time step $t'$. The dynamics between compartments are described by the following system of ordinary differential equations with given initial conditions.

$$\begin{aligned}
\frac{dS(t')}{dt'} &= \beta - (\nu + FOI_s(t))\ S(t') \\
\frac{dE(t')}{dt'} &= FOI_s(t)\ S(t') - (\nu + \tau)\ E(t') \\
\frac{dI(t')}{dt'} &= \tau\ E(t') - \nu_I\ I(t') \\
\frac{dC(t')}{dt'} &= \lambda\ I(t') - \gamma\ C(t')
\end{aligned}$$

Newly born snails are susceptible to schistosomiasis infection. Susceptible snails reproduce with a birth rate $\beta(t')$, according to a logistic growth due to competition for resources:

$$\beta(t') = \beta_0\ \left(1 - \frac{N_s(t')}{K}\right)\ (S(t') + E(t'))$$

Here, $\beta_0$ is the maximum reproduction rate in absence of competition, $N_s(t')$ the total snail population size, and $K$ the carrying capacity. $K$ is tuned to parameterise a mild or strong degree of snail-level regulating mechanism. Only susceptible $S(t')$ and exposed $E(t')$ snails are assumed to contribute to reproduction, due to castration upon parasitic infection.

Susceptible snails can encounter invasion of miracidia released in the water environment after egg excretion by human hosts and become exposed according to a linear force of infection $FOI_s(t) = \eta\ M(t)$ where $\eta$ is the transmission parameter on snails. Since the miracidial input $M(t)$ is a quantity obtained by the ABM simulation, the $FOI_s(t)$ is considered a constant parameter within the time horizon of the compartmental module.

Miracidia live in exposed snails for a maturation period $h$, during which they mature and asexually reproduce into cercariae. Susceptible and exposed snails undergo natural mortality, according to a mortality rate $\nu$. When the parasites are patent, the hosting snails move to the infected stage $I(t')$ with a rate $\tau = \frac{1}{h}$. Infected snails experience increased mortality ($\nu_I$) and castration. Infected snails shed cercariae ($C(t')$) into the environment, according to a cercarial per capita production rate $\lambda$. Cercariae can naturally die with a rate $\gamma$ before infecting humans.

At each ABM-time step $t$, the SEIC model is run for a period equal to $h$, with the aim to approximate the miracidial maturation in the intermediate host. The state $(S(h), E(h), I(h))$ is then set as initial conditions for the ODE system at the following ABM-time step $t+1$. Similarly, the cercarial amount is updated as $C(t+1) := C(h)$ and it will contribute to the definition of the $FOI_H(t+1)$.

#### Mass drug administration

SchiSTOP allows the incorporation of control activities such as Mass drug administration (MDA). MDA with praziquantel is implemented as a process that causes the death of adult worms (with a certain probability) in the treated individuals. Several parameters can be varied to reflect different options of treatment campaigns and efficacy.

- Length, namely how many years the MDA campaign last for
- Frequency, in terms of how often MDA campaigns implemented e.g. every second year, every year, twice a year

- Target population eligible for the treatment, e.g. School-aged children only or the whole community
- Coverage: portion of the target population actually reached by the treatment
- Target population systematically untreated: fraction of the target population that is not reached by MDA over consecutive rounds
- Drug efficacy: fraction of killed adult worms (pairs) per each human host

We assume that 100% of the target population adheres to the MDA programme. It is important to note that only adult worms die with praziquantel. Killed worms contribute to the amount of accumulated dead worms that in turn act as a trigger for the development of anti-reinfection immunity in our model.

Table 1: **Parameters employed in the model.**

| **Parameter** | **Value** | **Source** |
|---|---|---|
| **Human demography** | | |
| Birth rate | 36.5 | [2] |
| Emigration rate | 20 | Tuned |
| Eligible age group for migration [years old] | $[5 - 55]$ | [6] |
| Death probabilities by age | $[0 - 1]$ | [2] |
| **Parasite life within human host** | | |
| Aggregation of worms ($k_w > 0$) | - | Varying |
| Transmission parameter on humans ($\zeta > 0$) | - | Varying |
| Age specific relative exposures | *Model-based*: Piece-wise constant. (0.032, 0.61, 1, 0.06) for ages (0-4, 5-9, 10-15, 16+).<br>*Based on water contacts*: Piece-wise linear. (0, 0.62, 1, 0.51, 0.51) for ages (0, 5, 15, 40+). | [7–10] |
| External force of infection | Value = 1 worm<br>Duration = 3 years | Assumption |
| Average lifespan of worms within the human host [months] | 60 | [11] |

Continued on next page

Table 1: **Parameters employed in the model.** (Continued)

| **Parameter** | **Value** | **Source** |
|---|---|---|
| Pre-patent period [months] | 3 | [11] |
| **Egg production** | | |
| Expected number of eggs per sample ($\alpha > 0$) [eggs/worm pair/sample] | [0.12 - 0.14] | [12] and **Methods** main text |
| Density dependence in egg production ($z$) [/ female worm] | (Absent) 0, (Mild) 0.0005, (Strong) 0.0007 | [4] and **Methods** main text |
| Daily grams of stool produced by each human individual [gr] | 150 | [13] |
| Aggregation of observed egg counts ($k_e > 0$) | 0.87 | [13] |
| **Anti-reinfection immunity** | | |
| Immunity coefficient ($\alpha_{imm} > 0$) | (Absent) 0, (Mild) 0.0005, (Strong) 0.002 | [14] and **Methods** main text |
| **Snail population module** | | |
| Maximum reproduction rate ($\beta_0 > 0$) [1 / days] | 1 | [5] |
| Carrying capacity ($k > 0$) [number of snails] | (Absent) -, (Mild) 20000, (Strong) 10000 | Varying |
| Natural mortality of snails ($\nu > 0$) | $v = \frac{1}{100\ days}$ | [5] |
| Mortality of snails upon infection ($\nu_I > 0$) | $v = \frac{1}{30\ days}$ | [5] |
| Snail transmission parameter ($\eta > 0$) | - | Varying |

Continued on next page

Table 1: **Parameters employed in the model.** (Continued)

| **Parameter** | **Value** | **Source** |
|---|---|---|
| Worm maturation period within the snail ($h > 0$) [days] | 30 | [4, 15] |
| Cercarial production rate ($\lambda > 0$) [1/days] | 50 | [5] |
| Mortality rate of cercariae ($\gamma > 0$) [1/days] | 1 | [5] |
| **Mass drug administration** | | |
| Target population | $5 - 15$ or 2+ years old | Assumption |
| Duration | 10 years | Assumption |
| Frequency | Annual | Assumption |
| Coverage | 75% | [16] |
| Efficacy | 86% | [8] |
| Fraction systematically untreated | 5% of target population | [8] |

16. World Health Organization. WHO guideline on control and elimination of human schistosomiasis. Geneva: WHO; 2022.
