## Supplementary material for "Revisiting the impact of *Schistosoma mansoni* regulating mechanisms on transmission dynamics using SchiSTOP, a novel modelling framework": S2 Appendix

### Effectiveness of treatment and prevalence bounce-back

#### Prevalence timelines in SAC during and after treatment.

For each choice of the age-exposure function and endemicity setting (titles), single panels refer to a fixed combination of the assumptions of human-level regulation (columns) and snail-level regulation (rows). A single panel shows the infection prevalence in school-aged children on the y-axis (mean of 100 stochastic realizations of the model, single runs as shaded lines) by round of treatment on the x-axis. The prevalence timelines are displayed by varying the degree of worm-level regulation (colours). For all scenarios, treatment is annually administered to 5-15 years old individuals with 10 repeated rounds, a coverage of 75%, 5% of target population systematically untreated, and a drug efficacy of 86%.

1. **Assumption for the age-exposure function: "Model-based function"**

Low endemicity setting

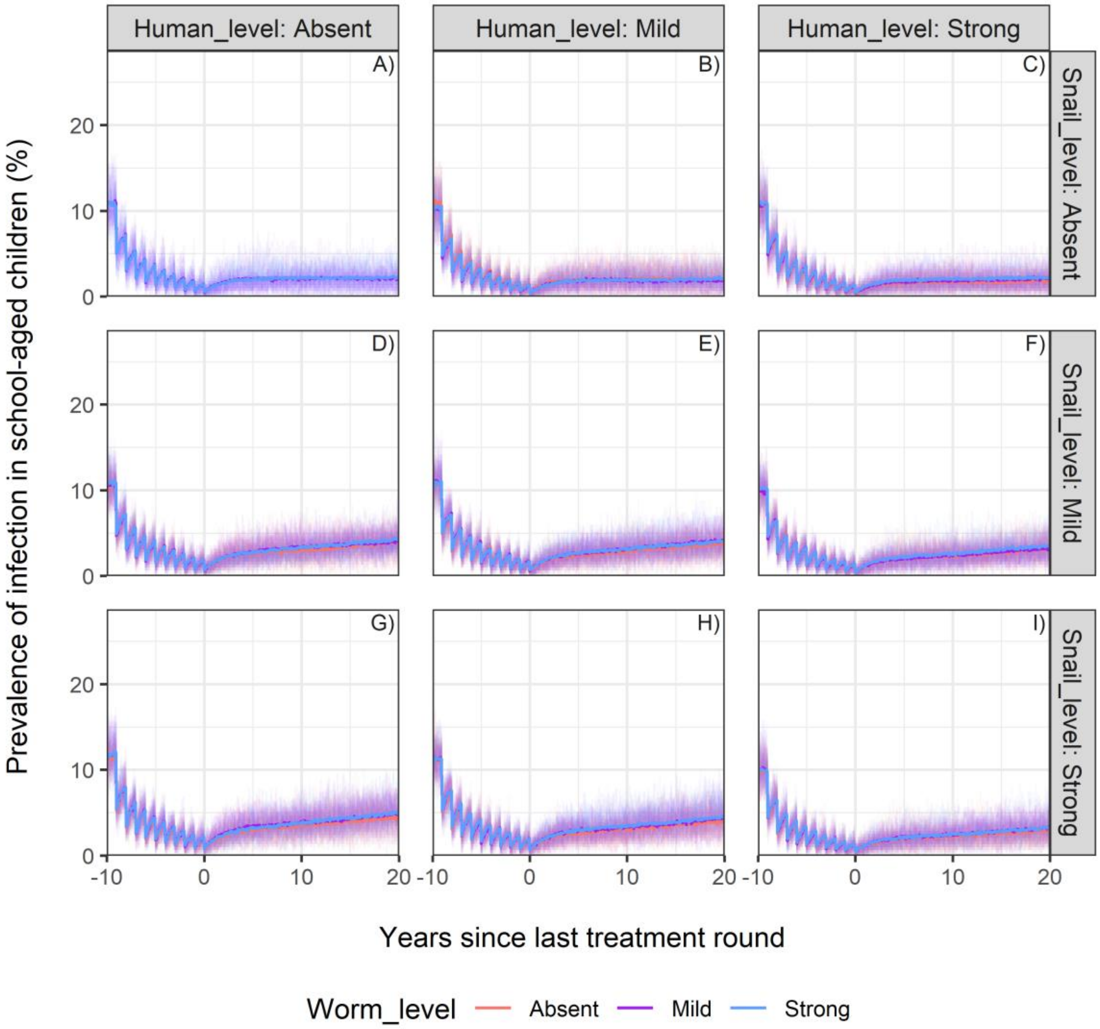

Moderate endemicity setting

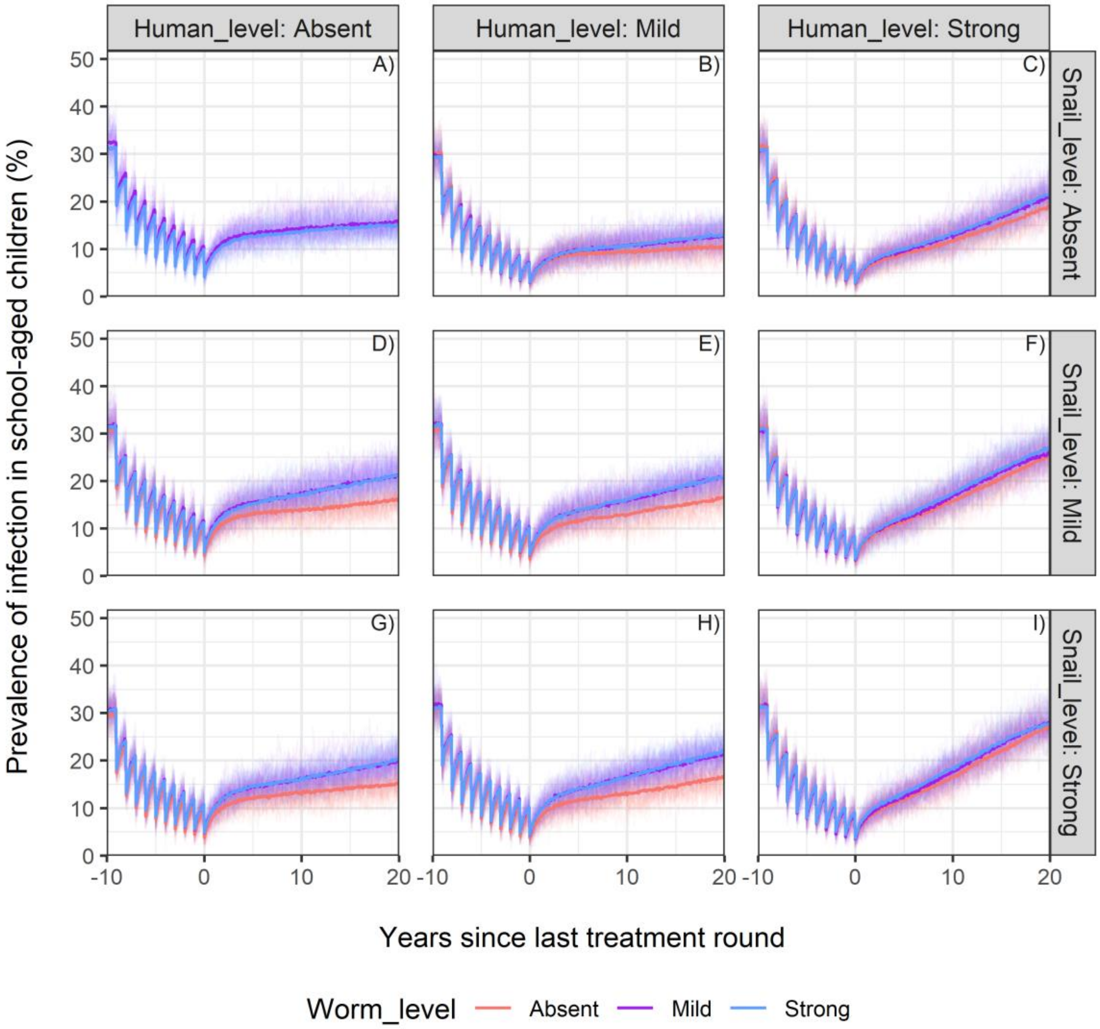

High endemicity setting

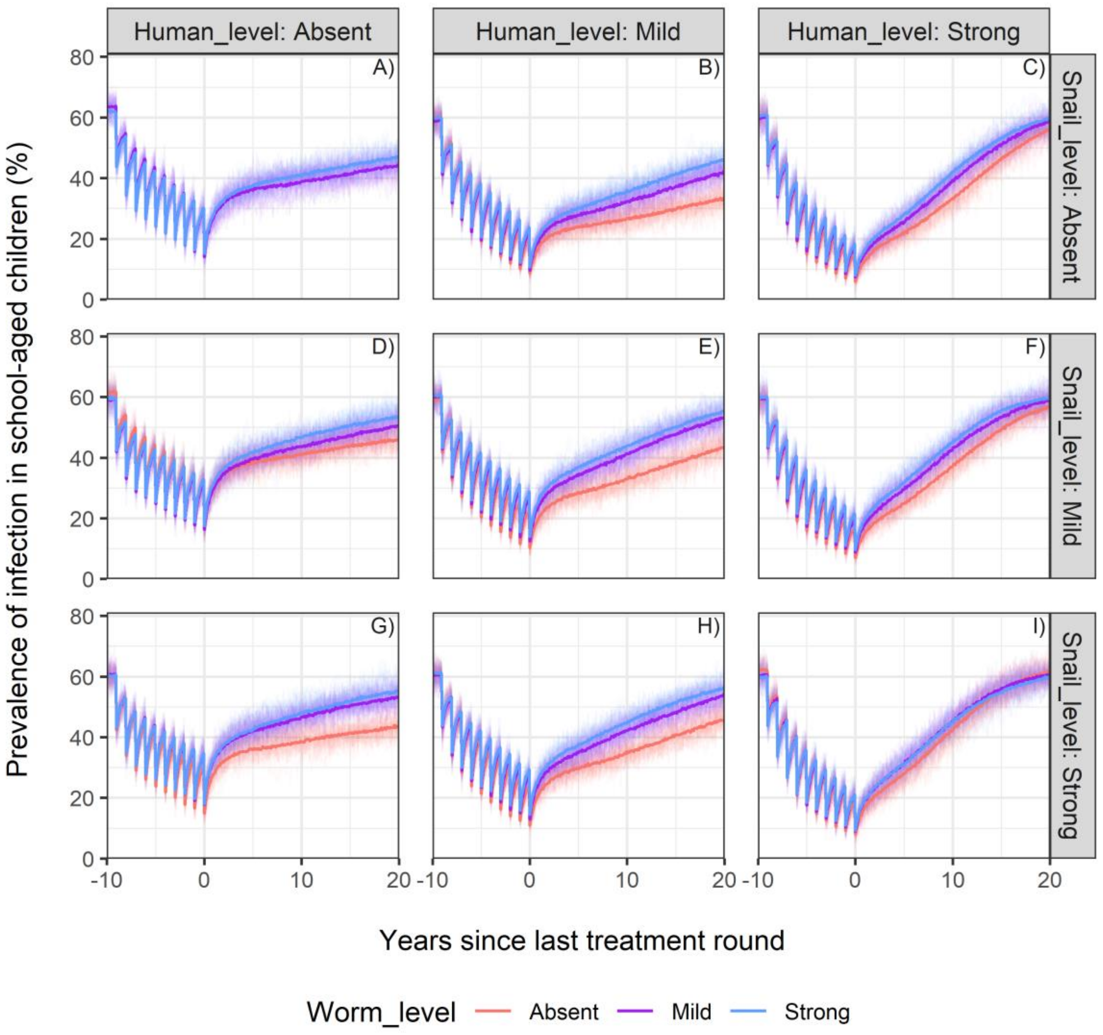

### 2. Assumption for the age-exposure function: "Based on water contacts"

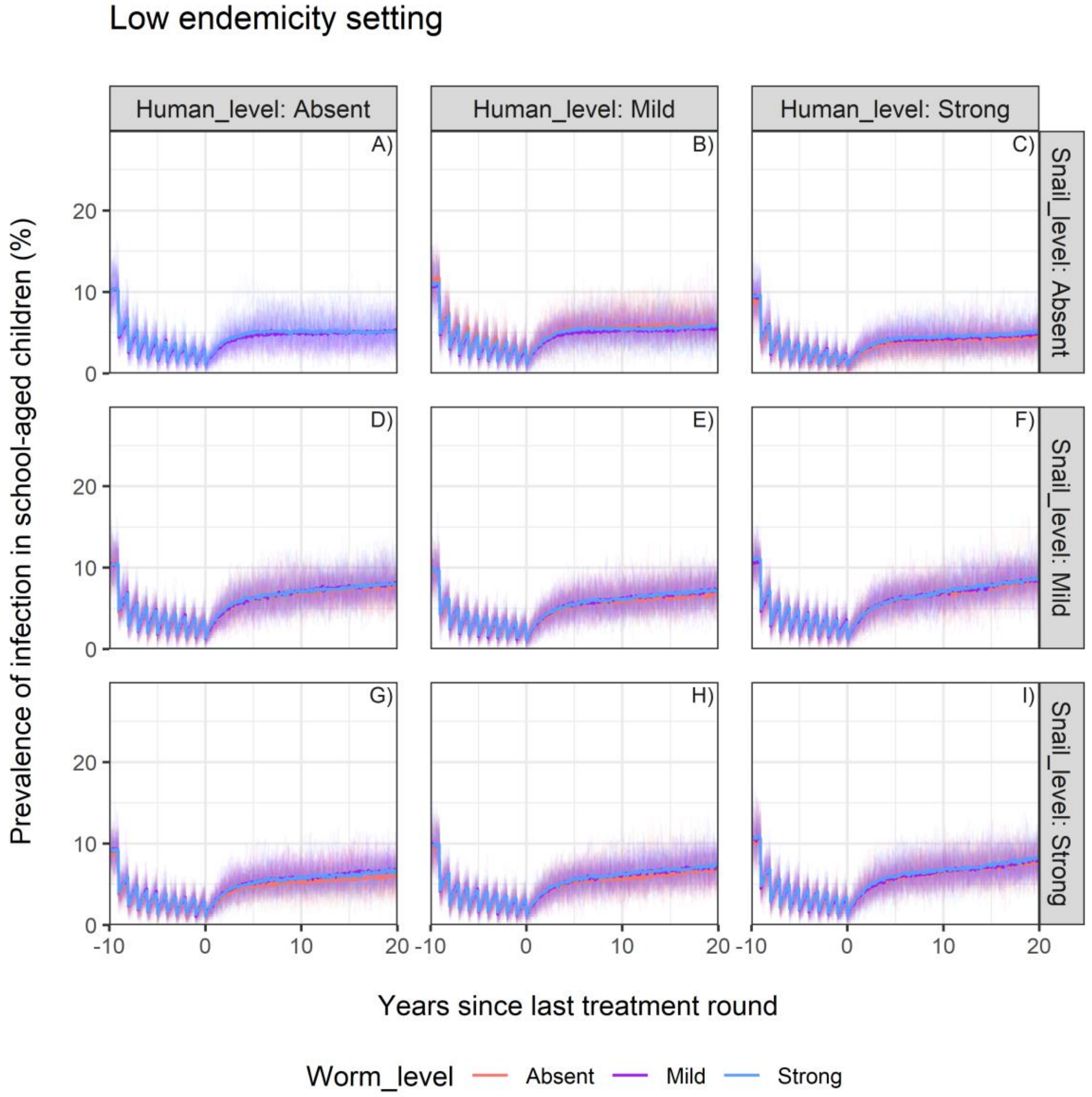

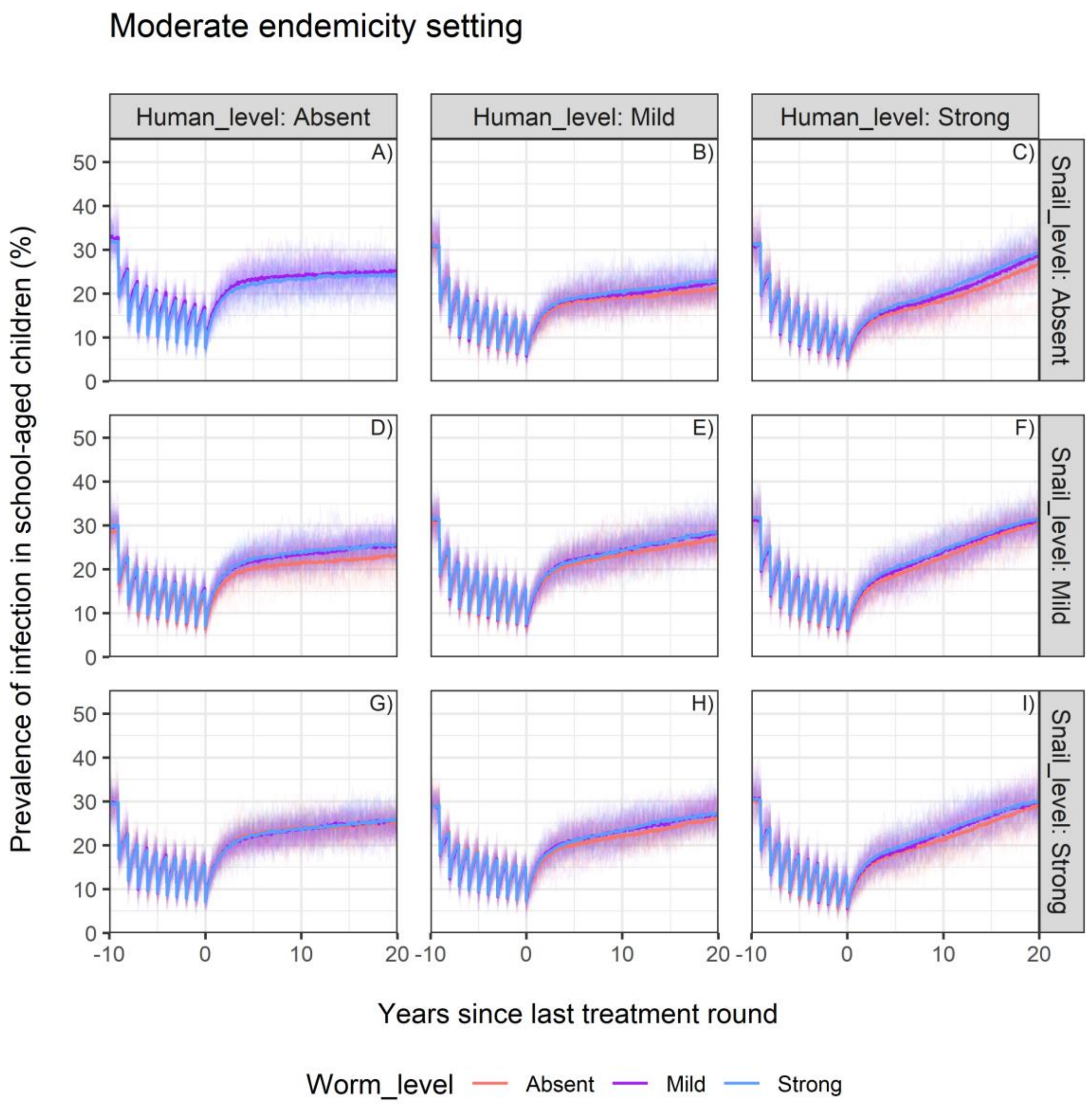

Moderate endemicity setting
Human_level: Absent
Human_level: Mild
Human_level: Strong
Snail_level: Absent
Snail_level: Mild
Snail_level: Strong
A)
B)
C)
D)
E)
F)
G)
H)
I)
Prevalence of infection in school-aged children (%)
Years since last treatment round
Worm_level
Absent
Mild
Strong

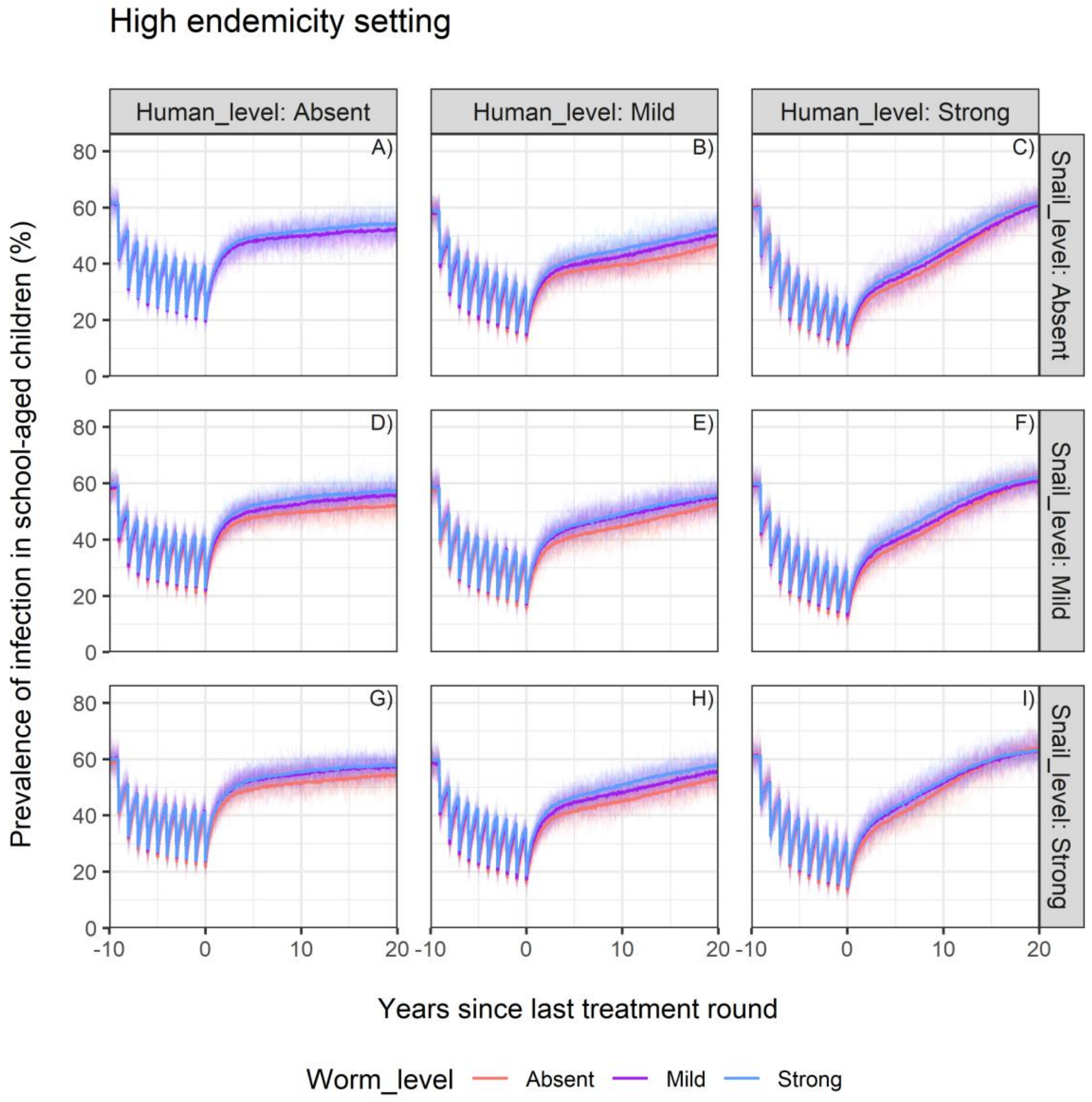
High endemicity setting
Human_level: Absent
Human_level: Mild
Human_level: Strong
Snail_level: Absent
Snail_level: Mild
Snail_level: Strong
A)
B)
C)
D)
E)
F)
G)
H)
I)
Prevalence of infection in school-aged children (%)
Years since last treatment round
0
20
40
60
80
-10
0
10
20
Worm_level
Absent
Mild
Strong
